# Proteasome inhibition enhances myeloma oncolytic reovirus therapy by suppressing monocytic anti-viral immune responses

**DOI:** 10.1101/2022.03.29.22272857

**Authors:** Ada Alice Dona, Enrico Caserta, Mahmoud Singer, Theophilus Tandoh, Lokesh Nigam, Janet Winchester, Arnab Chowdhury, Yinghui Zhu, Mariam Murtadha, Alex Pozhitkov, James F Sanchez, Hawa Vahed, Matt Coffey, Guido Marcucci, Amrita Krishnan, Gerard Nuovo, Douglas W. Sborov, Craig C Hofmeister, Flavia Pichiorri

## Abstract

Reovirus is an oncolytic virus with natural tropism for cancer cells. We previously showed that reovirus intravenous administration in myeloma patients was safe, but disease control associated with viral replication in the cancer cells was not observed. Here we show that *ex vivo* proteasome inhibitors (PIs) potentiate reovirus replication in circulating classical monocytes, increasing viral delivery to myeloma cells. We found that the anti-viral signals in monocytes primarily rely on the NF-kB activation and that this effect is impaired by the addition of PIs. Conversely, PIs improved reovirus-induced monocyte and T cell activation against cancer cells. Based on these preclinical data, we conducted a phase 1b trial of the reovirus Pelareorep together with the PI carfilzomib in 13 heavily pretreated bortezomib-resistant MM patients. Objective responses associated with reovirus active replication in MM cells, T cell activation and monocytic expansion were noted in 70% of patients.

## INTRODUCTION

Oncolytic viruses preferentially target and kill cancer cells without destroying normal cells. Talimogene laherparepvec (TVEC), the first FDA-approved oncolytic virotherapy, is a modified herpes simplex virus-1 (HSV-1) that encodes granulocyte-macrophage colony-stimulating factor (GM-CSF) and is now used for the treatment of metastatic melanoma *(1)*. Although oncolytic viruses for patients with solid tumors have primarily been injected in a specific tumor mass, successful treatment of hematologic malignancy with an oncolytic virus requires intravenous infusion to transport the virus through the bloodstream to bone marrow (BM) or extramedullary sites of disease to infect cancer cells and lead to cancer cell death, either through direct cytolysis or by engaging the immune microenvironment.

Reovirus Serotype 3 - Dearing Strain (RV) is a ubiquitous, non-enveloped, double-stranded RNA virus *(2)* that can cause mild gastroenteritis, coughing, and various flu-like symptoms *(3)*; it has been implicated in the initiation of celiac disease by promoting the loss of tolerance to dietary antigens *(4)*. The therapeutic form of RV, Pelareorep, was granted orphan drug status from the FDA and has been infused intravenously in numerous clinical trials. It enacts anti-neoplastic effects through both apoptotic and non-apoptotic mechanisms *(5)*. Due to RV’s natural tropism for transformed cells and its relatively non-pathogenic profile, it is considered an ideal non-engineered virus for oncolytic therapy *(6, 7)*.

RV entry into cells requires sequential binding, first to extracellular sialic acid followed by engagement of junctional adhesion molecule A (JAM-A) *(8-10)*, a receptor that is expressed on the surface of myeloma (MM) cells *(11, 12)*. RV is capable of selectively replicating in transformed cells with impaired intracellular antiviral responses, leading to selective anti-tumor activity *(13, 14)*. Preclinical investigation of RV for the treatment of MM has demonstrated significant anti-MM effects *in vitro* and *in vivo* when used as a single agent and in combination with proteasome inhibitors (PIs) and histone deacetylase inhibitors *(15, 16)*, but proposed mechanisms associated with the anti-MM activity of RV in combination with these agents have been unclear. Our initial trial, the first investigation of RV in any hematologic malignancy, used intravenous RV as a single agent in relapsed MM *(17)*. This trial showed that RV was able to reach MM cells in the marrow; however, it was unable to induce a productive infection or confer disease control through cytolytic killing of the cancer cells. Recently published preclinical data in an immune competent MM mouse model also show that intravenous RV did not replicate within BM myeloma cells, but instead led to a significant immune reaction *(18)*.

Treatment with PIs (e.g., bortezomib [BTZ], carfilzomib [CFZ]), immunomodulatory drugs (e.g., lenalidomide, pomalidomide), CD38-directed antibodies (e.g. daratumumab and isatuximab), and dexamethasone is currently standard of care for MM patients *(19)*. One mechanism of action attributed to PIs is the killing of MM cells via inhibition of NF-κB activation, by blocking the proteasomal degradation of the main NF-κB inhibitor, IκB⍰ *(20)*. Previous data have shown that RV-induced MM cell killing is potentiated by BTZ, either as a direct effect against MM cells by increasing RV-induced endoplastic reticulum stress *(21)* or by enhancing permissivity of the viral replication in tumor-associated endothelial cells, leading to enhanced viral delivery to MM cells and thereby stimulating cytokine release *(22)*. However, molecular mechanisms behind this observation were not explored.

Here, using human primary samples and MM animal models, we investigated the mechanisms behind the ability of PIs to enhance the activity of RV (Pelareorep). We demonstrated that the immune-modulatory activity of PI positions it as an ideal therapeutic companion to enhance oncolytic viral therapy. We also report the results of the first two cohorts of a phase 1b trial of CFZ+Pelareorep along with correlative studies supporting the use of a PI in combination with oncolytic viral therapy.

## RESULTS

### PIs potentiate RV induced MM cell killing only with the involvement of the microenvironment

In line with previously published results *(22)*, our data show that PIs do not enhance RV viral replication in MM cell lines. Specifically, when MM cell lines with moderate (MM.1S, L363, H929) and high (RPMI-8226, U266) viral tropism *(12)* were treated with Pelareorep in combination with CFZ, we observed either decreased or unchanged levels of RNA genome **(Fig. 1A)** and sigma non-structural capsid protein (σ-NS), compared to cells treated with Pelareorep alone **(Fig. 1B, C)**. Similar effects were observed when the PI BTZ was used **(Supp. Fig. 1A, B, C)**. Additionally, our data show that at early timepoints (24-48hrs), PI treatment does not increase Pelareorep-induced apoptosis in all of the MM cell lines we tested **(Fig. 1D, E and Supp. Fig. 1D, E)**.

**Figure 1.**
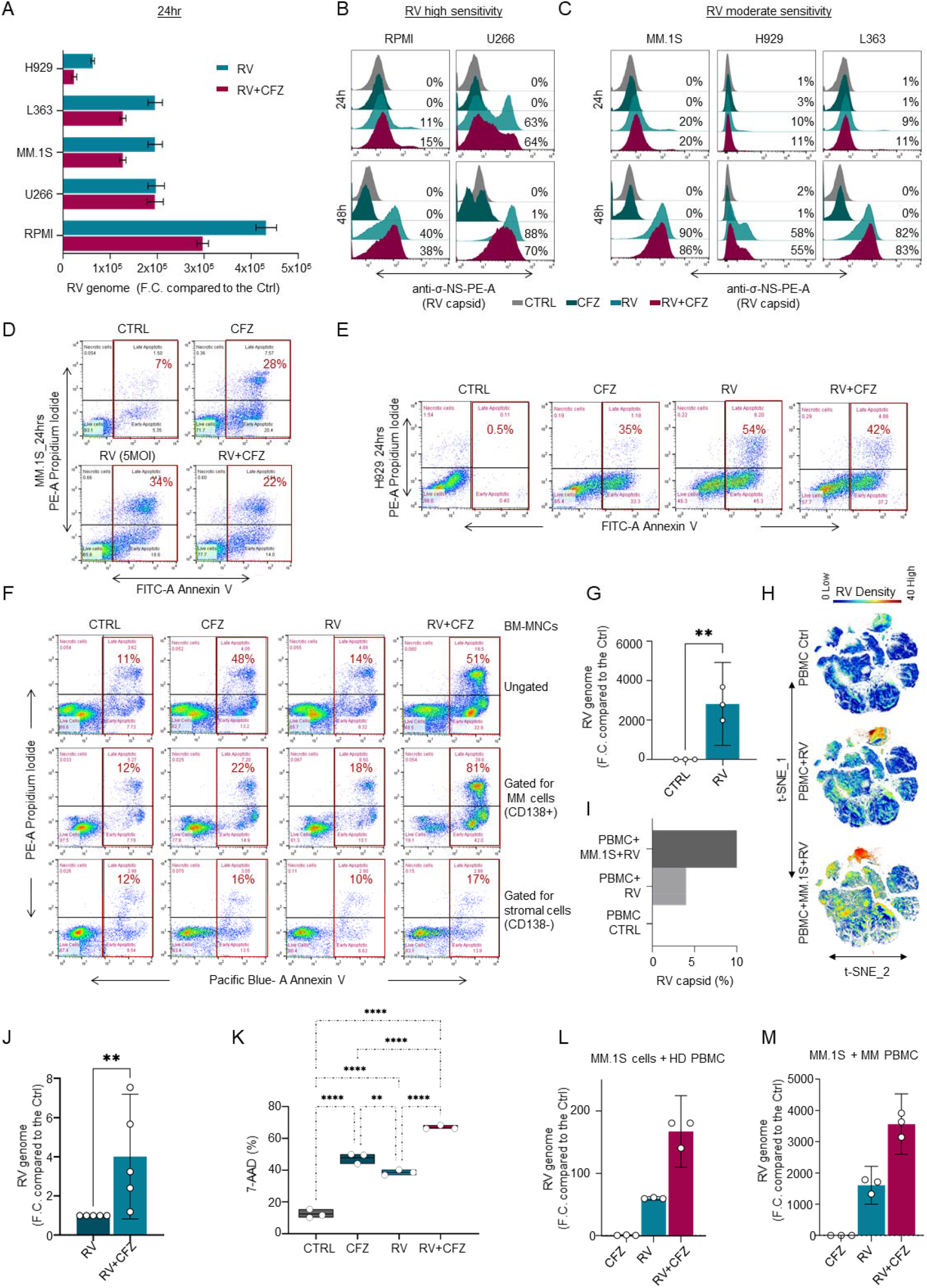
Proteasome inhibitor potentiates RV-induced MM cell killing only with the involvement of the microenvironment. **(A)** MM cell lines (RPMI-8226, U266, MM1.S, L363, H929) were treated with CFZ (5 nM) and Pelareorep (5 MOI) for 24hrs to assess the RV genome (mRNA) expression by q-RT-PCR; (**B-C)** Offset histograms showing sigma non-structural capsid protein (σ-NS) on cell lines with high (RPMI-8226, U266) and moderate (MM1.S, L363, H929) sensitivity to Pelareorep infection treated with CFZ (5 nM) and infected with Pelareorep (5 MOI) for 24 and 48hrs; (**D-E)** Representative flow cytometry plots using Annexin V-FITC/PI staining for apoptosis, showing the apoptotic rate (%) of MM1.S (D) and H929 (E) cells exposed to CFZ (5 nM) and RV (5 MOI) alone or in combination for 24hrs; (**F)** Primary CD138+ MM-PCs of a SMM patient were isolated from the CD138 (-) BM-MNCs, which were treated with CFZ (5 nM) and RV (10 MOI) alone or in combination overnight, washed, and cocultured with the autologous CD138+ MM cells for 24hrs to identify apoptotic cells (Annexin V +) and analyzed by flow cytometry gating on total BM-MNCs, CD138+ BM-PCs, and the CD138neg fractions; (**G)** RV capsid mRNA level expression in healthy donor (HD) PBMCs infected with RV (10 MOI) as fold change (F.C.) compare to the CTRL. Data are expressed as the mean ± SEM (n=3 HDs), normalized compared to control GAPDH; **p≤0.01; (**H)** A 34-Ab CyTOF panel was used to generate FCS files. Hierarchical clustering and statistical mapping was performed algorithmically via Cytobank© platform. vi-SNE analysis (iterations=4000, perplexity=50) is displayed as 2D plots using the resultant t-SNE_1 and t-SNE_2 dimensions. t-SNE heatmaps according to the density expression of RV capsid (Tb159Di) were gated for the total leukocytes from an HD PBMC infected or not with RV (10 MOI), or in Pelareorep-treated PBMCs to which we added for 1hr MM1.S cells that had been pre-infected for 24hrs; (**I)** Bar graph showing RV capsid signal intensity (%) from treatment conditions in (H); (**J)** q-RT-PCR showing RV genome expression in F.C. compared to the CTRL in HD PBMCs infected with RV (5 MOI) and/or CFZ (2.5 nM) for 24hrs. Data are expressed as the mean ± SEM (n=5 HDs), normalized compared to control GAPDH; **p≤0.01; (**K)** Flow cytometry-based killing assay using PBMCs isolated from HD, treated for 30 minutes with or without CFZ, then infected overnight with or without RV, washed, and co-cultured (8:1) with Gfp+ MM.1S for 12hrs. Data are expressed as the mean ± SEM in triplicates in two different biological systems (n=2 HDs); ****p≤ 0.0001; **p≤0.01; (**L-M)** q-RT-PCR of the viral genome expression of PBMCs isolated from an HD (L) or an MM patient (M), treated or not with RV (5 MOI) and CFZ (2.5 nM), and co-cultured with MM1.S cells for 12hrs. Data are expressed as the mean ± SEM in triplicates.

We then investigated whether PIs could instead increase Pelareorep-induced killing of MM cells in the presence of the tumor microenvironment, independently of its direct anti-MM activity, in both an animal model, as recently published *(22)*, and in the *ex vivo* setting. CD138+ MM cells were removed from the BM cells (BM-MNCs), which includes both stromal and immune cells, that were obtained from a patient with smoldering MM. The resulting CD138(-) BM-MNCs were then treated overnight with CFZ or Pelareorep alone or in combination, washed with PBS, and then co-cultured with the autologous CD138+MM cells for 24hrs. Our data show increased cell death in the CD138+ fraction (MM cells) when the CD138(-) BM-MNCs were pre-treated with PI+Pelareorep, compared to the single agents **(Fig. 1F)**. Of note, no increased cell death was observed in the CD138(-) fraction in the same experimental conditions (**Fig. 1F**). MM adhesion to the BM stromal cells displayed resistance to RV infection and induced oncolysis (**Supp. Fig. 1F**), excluding that these cells may contribute to the enhancement of RV-induced MM cell killing activity by PIs. Because it has been reported that, despite the presence of neutralizing antibodies, replication-competent RV can be recovered from peripheral blood mononuclear cells (PBMCs) but not plasma obtained from colorectal cancer patients treated intravenously with a single dose of RV *(23)*, we decided to investigate whether the immune environment would instead be responsible for supporting RV replication and subsequent MM cell infection. When healthy human PBMCs were treated with Pelareorep for 24hrs, an increase in the viral genome (p=0.004, n=3 donors) and capsid were observed **(Fig. 1G, I)**, supporting that a limited viral replication can be detected even in non-cancer cells. Consistent with these data, single cell mass cytometry (CyTOF) analysis show a localized expression of the inner viral capsid protein σ-NS, whose expression is associated with active viral replication *(12, 24)*, in PBMCs treated *ex vivo* with Pelareorep for 24 hours (**Fig. 1H, I and Supp. Fig. 1G**). A greater increase in capsid expression was found when MM.1S cells were pre-treated for 24 hrs with Pelareorep and added to Pelareorep-treated PBMCs for 1hr (**Fig. 1H, I and Supp. Fig. 1H)**, supporting the specificity of intracellular capsid detection by CyTOF.

These findings prompted us to investigate whether PIs could indeed increase viral replication in PBMCs. Our data show that the addition of CFZ significantly increased RV replication in healthy donor PBMCs (n=5) after 24hrs of treatment (p=0.008) **(Fig. 1J)**, an effect that was also observed at earlier timepoints **(Supp. Fig. 1I)**. Increased viral replication in PI-treated healthy donor PBMCs also enhanced subsequent viral delivery and killing of MM cells. Specifically, when PBMCs were treated with CFZ+Pelareorep for 24hrs, then washed and co-cultured for 12hrs with untreated MM cells, previously unexposed to the virus, at the ratio 8 (PBMCs):1 (MM.1S GFP+ cells), the PI-pretreated PBMC fraction strongly enhanced RV-induced anti-MM activity (p<0.0001), compared to that of RV alone **(Fig. 1K)**, an effect that was associated with increased levels of viral genome in MM cells (MM.1S Gfp+) **(Fig. 1L)**. Increased levels of viral genome were also observed in MM cells co-cultured with PBMCs obtained from MM patients **(Fig. 1M)**, supporting that the effect of PIs in increasing viral replication is a general phenomenon.

### Proteasome inhibitor-enhanced viral replication requires monocytes

Because our data suggested that PIs (CFZ and BTZ) improve RV replication in PBMCs and subsequent infection and killing of cancer cells, we investigated which immune compartment is responsible for this effect.

Twenty-two different immune compartments including CD4+ and CD8+ T cell subsets (naïve, central memory [CM], effector memory [EM] and terminally differentiated [TEMRA]), natural killer T cells (NKT), classical phagocytic and non-classical monocytes, and NK cells **(Supp. Table 1)** were interrogated for the presence of viral capsid protein (σ-NS) using CyTOF. After 24 hours of Pelareorep treatment, the RV capsid was mainly found in the monocyte compartment (CD45+CD20-CD3-CD56-CD14+HLA-DR+) **(Supp. Fig. 2A, B)**. Significant capsid accumulation in the CD14+ fraction, compared to the matched CD14(-) fraction, was observed in all donor PBMCs we tested (p=0.03, n=4) **(Fig. 2A)**. Cell cluster visualization self-organizing map (FlowSOM) **(Fig. 2B)** and t-distributed stochastic neighbor embedding (t-SNE) heatmaps (**Fig. 2C)** showed that the σ-NS signal (red) was primarily observed in the Classical monocytes (phagocytic). Specifically, we found active viral replication at 24hrs in 10% +/- 3.05% of the phagocytic monocytes, but almost no replication was observed in both Intermediate and Non-Classical monocytes **(Fig. 2B, C)**.

**Figure 2.**
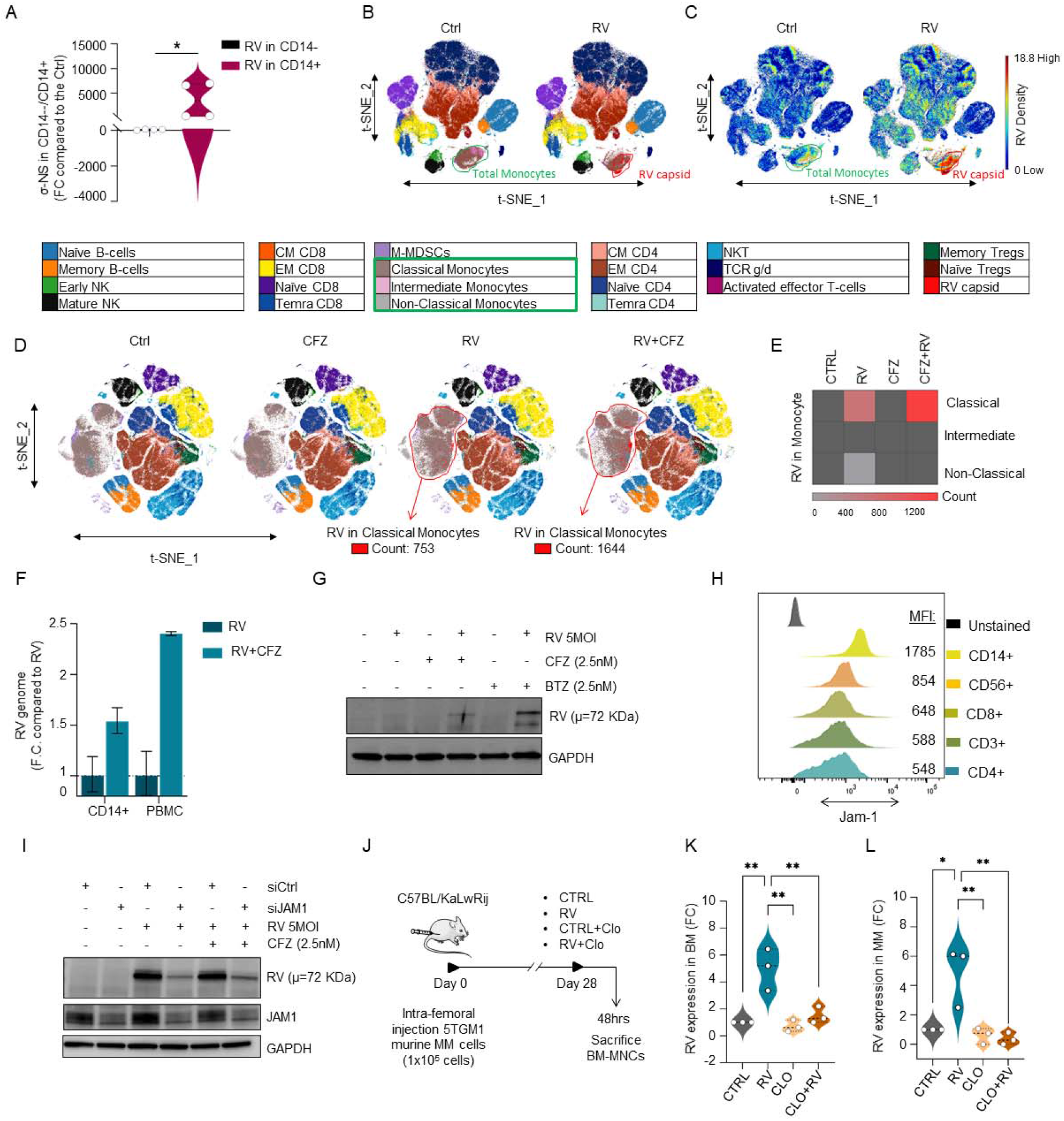
Proteasome inhibitor-enhanced viral replication requires monocytes. **(A)** Violin-plot showing (σ-NS) signal in 4 different HDs analyzed by CyTOF *p≤0.05; (**B-C)** 34-Ab CyTOF panel used to generate FCS files. Hierarchical clustering and statistical mapping performed algorithmically via the Cytobank© platform. vi-SNE analysis (iterations=3000, perplexity=100) displayed in 2D plots using the resultant t-SNE 1 and t-SNE2 dimensions. High-fidelity FlowSOM (“self-organizing map”) (metacluster=10 and cluster=100) based on vi-SNE 2D plots showing 22 different immune compartments in total HD-PBMCs infected or not with RV (10 MOI). Red signal shows RV capsid (σ-NS) in monocytes (B. t-SNE heatmap highlighting density expression of selected RV capsid (σ-NS) signal (C; **D-E)** CyTOF high-fidelity FlowSOM in HD-PBMCs infected or not with RV (10 MOI) alone or in combination with CFZ (2.5 nM), showing increased RV capsid (σ-NS) detection in Classical Monocytes after RV+CFZ co-treatment (RV count=753, RV+CFZ=1644) (D), and heatmap graphical representation of RV absolute count detection in the different monocyte subsets (E); (**F)** q-RT-PCR for the viral genome expression of RV-infected HD-PBMCs or isolated HD-CD14+ population after 24hrs, normalized to control GAPDH and expressed as the mean ± SEM of triplicates in fold change (FC) compared to RV alone; (**G)** Western Blot analysis of (σ-NS) viral protein in HD CD14+ selected population after 8hrs of PIs (CFZ and BTZ 2.5 nM) and RV treatments alone or in combination; (**H)** Offset histograms showing JAM-1 flow cytometry detection in different immune subsets (CD14+, CD56+, CD8+, CD4+, CD3+). The experiment was repeated in triplicate; (**I)** Western blot analysis on THP-1 showing RV (σ-NS) protein detection after specific JAM-1 knockdown; (**J)** Schematic representation of mice experiment: 12 immune competent myeloma mice (C57BL/KaLwRij) were injected intra-femorally with 1×10^5^ 5TGM1 murine MM cells and treated with 150 mg/kg clodronate-liposome for monocytes-macrophages depletion or control, then with or without intravenous injection of RV (5 × 10^8^ TCID_50_) for 48hrs; (**K-L)** Violin plots showing bone marrow RV (σ-NS) capsid formation (**p≤0.01) (K) of treated mice and in MM-CD138+ cells (**p≤0.01 and *p≤0.05) (L), analyzed by flow cytometry.

We then investigated whether PIs could increase RV replication in the monocytic fraction. When PBMCs were treated with Pelareorep alone or in combination with CFZ, viral replication occurred exclusively in the Classical monocyte population, with more replication evident following combination treatment (absolute count: 1528 [RV and PI] vs 704 [RV alone]) as shown by FlowSOM (**Fig. 2D)**, heatmap graphical representation from CyTOF (**Fig. 2E and Supp. Fig. 2C, D)**, and flow cytometry analysis **(Supp. Fig. 2E)**. Analysis conducted in primary CD14+ selected populations from healthy donor PBMCs confirmed that the addition of PI to RV-infected cells enhanced genome infection **(Fig. 2F)** and replication based on detection of σ-NS **(Fig. 2G)**. An increase in RV genome replication upon CFZ treatment was not noted in all other major immune subsets, including B, T and NK cells **(Supp. Fig. 2F)**.

Because we found active viral replication in the CD14+ fraction, we tested whether the junctional adhesion molecule 1 (JAM-1) receptor *(9)* mediates RV entry in this cellular subset. Flow cytometry analysis on different immune cell subsets in healthy donor PBMCs revealed high JAM-1 expression on the surface of the monocyte population **(Fig. 2H and Supp. Fig. 2F)**. Following knockdown of JAM-1 expression in a monocytic-like cell line (THP-1), viral replication was strongly impaired **(Fig. 2I and Supp. Fig. 2G, H, I)**.

To assess whether the CD14+ fraction is critical for RV delivery to the cancer cells *in vivo*, we induced monocyte/macrophage depletion in an immunocompetent MM animal model using clodronate liposomes (Clo). Specifically, 1×10^5^ murine MM 5-TGM1 cells were intrafemorally injected into syngeneic C57BL/KaLwRij mice. After 28 days, mice were randomized to receive clodronate liposomes (Clo) plus RV (1×10^7^ termination of the 50% tissue culture infectious dose [TCID_50_]) or control liposomes (CTRL) plus RV (1×10^7^ TCID_50_). Mice treated with only Clo or control were also included as internal controls (**Fig. 2J)**. After 48hrs, RV capsid formation by flow cytometry in BM-MNCs and in MM-CD138+ cells was significantly lower in the Clo+RV treated group compared to that in the control+RV group (p=0.004) **(Fig. 2K, L and Supp. Fig. 2J)**.

### CFZ stimulates immune activation but impairs the monocyte-mediated antiviral response

Oncolytic viruses induce an antiviral immune response *(25)* that is often accompanied by nuclear factor kappa B (NF-κB) activation *(26)* and dysregulated release of inflammatory cytokines from monocytes to block active viral infection *(27)*. For this reason, we investigated whether Pelareorep could induce NF-κB activation in monocytes. Immunofluorescence analysis at both 2 and 4 hours after RV infection of a monocytic cell line (THP-1) showed a significant increase in p65 nuclear translocation compared to that in the RV-untreated cells (p<0.0001) **(Fig. 3A, B)**, an effect that was similar to the one observed in cells treated with the specific NF-κB activator TNF-α, **(Fig. 3A, B and Supp. Fig. 3A, B)**. A significant decrease in p65 nuclear translocation was observed when CFZ was added to RV-treated cells (p<0.0001), supporting that PI impairs NF-kB activation upon RV infection.

**Figure 3.**
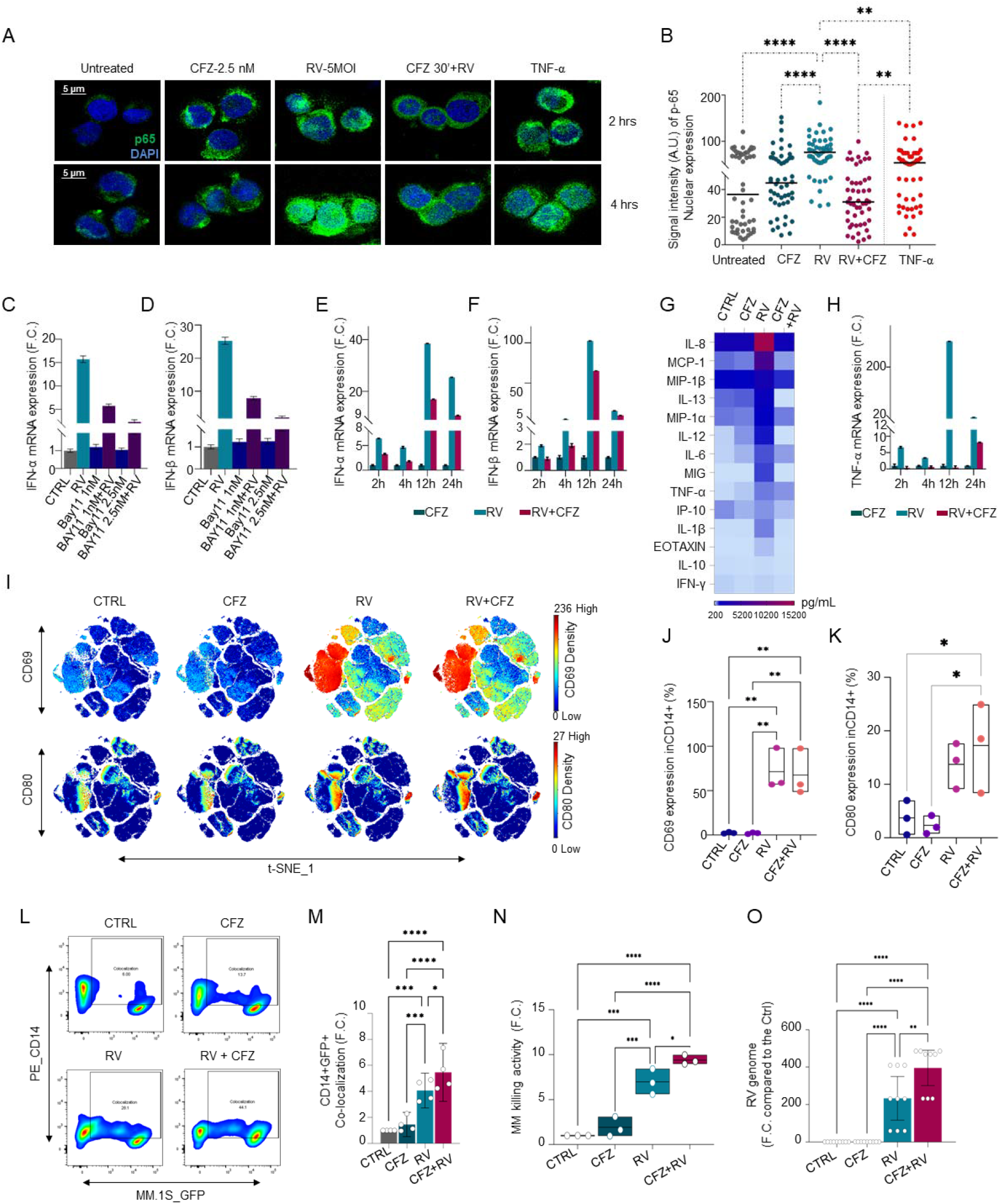
CFZ stimulates immune activation but impairs the monocyte-mediated antiviral response. **(A)** Representative merge of immunofluorescence fields showing p65 staining (green) as indicated and DAPI staining for nucleic acids (blue) in THP-1 cells treated with CFZ for 30 min, then infected or not with RV (5 MOI) for 2 or 4hrs. TNF-α (50 ng/ml) was used as a positive internal control showing p65 nuclear translocation; (**B)** Imaging-based quantification up to 2hrs of fluorescence staining intensity vs pixel position of 50 representative cells. Comparisons among groups were performed by one-way ANOVA: **≤0.01, ****p≤0.0001; (**C-D)** Histograms showing IFN-α and IFN-β mRNA expression of RV infected (5 MOI) or non-infected HD-PBMCs and treated with different concentration (1 or 2.5nM) of Bay11 for 24h and normalized to control GAPDH. Data represent the mean ± s.d. expressed in F.C. compared to the control; (**E-F)** q-RT-PCR showing IFNs type I (IFN-α and IFN-β) induction after RV (5 MOI) and CFZ (2.5 nM) treatments in HD PBMCs at different time points (2-4-12-24hrs). Data are normalized to control GAPDH and expressed as mean ± s.d.in F.C. compared to the control; (**G)** Heatmap of multiplex cytokine profile performed on supernatant from PBMCs from an HD treated for 4hrs with CFZ, RV or both, showing 14 out 22 of the analyzed cytokines; (**H)** Validation of TNF-α levels at 2-4-12-24hrs. Data represent the mean ± s.d. in F.C. compared to the control; (**I)** Mass cytometry t-SNE heatmaps showing CD69 and CD80 expression in HD PBMCs with or without RV infection (10 MOI) for 24hrs; (**J-K)** Box and whiskers plot for CD69 (J) and CD80 (K) relative expression. Comparisons among groups were performed by one-way ANOVA: for CD69 **p≤0.01; for CD80 *p≤0.05; (**L-M)** Flow cytometry showing increased CD14+ co-localization with MM1.S GFP+ cells after overnight incubation with RV (5 MOI) and/or CFZ (2.5 nM) treatments (L), and bar graph, n= 4 biological systems ****p≤0.0001, ***p≤0.001, *p≤0.05 (M); (**N)** Box and whiskers plot showing MM killing activity in n=3 biological replicates ****p≤ 0.0001, ***p≤0.001, **p≤0.01; (**O)** Relative mRNA expression level of RV genome in purified HD CD14+ cells treated with CFZ (2.5 nM) and/or RV (5 MOI), n=3 biological triplicates, ****p≤0.0001, ***p≤0.001, **p≤0.01.

Since our data suggested that CFZ increases RV replication in circulating monocytic cells **(Fig. 2F)**, we investigated whether blocking NF-kB activation could impair the expression of the anti-viral IFN-I response. Our data show that RV infection of PBMCs increased IFN-α and IFN-β expression, an effect that was reverted by the addition of the NF-kB specific inhibitor Bay-11 **(Fig. 3C, D)**. As expected, IFN-α and IFN-β expression were also significantly impaired in CFZ+RV treated cells compared to that from RV alone at different timepoints **(Fig. 3E, F and Supp. Fig. 3C, D)**, an effect that was observed exclusively in the monocytic fraction (CD14+ population) **(Supp. Fig. 3E, F)**.

Cytokine array analysis confirmed that RV-treated PBMCs for 4 hours released pro-inflammatory cytokines (IL-8, IL-10, IL-12, IL-13, and TNF-α) and chemokines (MIP-1 α and β, MIG, IL-8, MCP-1), an effect that was almost completely abrogated by adding CFZ **(Fig. 3G and Supp. Fig. 3G)**. q-RT-PCR analysis conducted at earlier and at later timepoints of RV treatment (2-24 hours) also confirms that the addition of CFZ significantly arrested TNF-α transcriptional activation **(Fig. 3H and Supp. Fig. 3H)**.

In-depth clustering CyTOF analysis revealed that RV infection induced significant upregulation of the early activation marker CD69 and T cell costimulatory receptor CD80 in the monocytic fraction of human PBMCs **(Fig. 3I, J, K and Supp. Fig. 3I, J)**. Addition of CFZ to RV did not affect the expression of either the monocytic activation marker CD69 or the differentiation marker CD80 **(Fig. 3I, J, K)**. The increase in T cell co-stimulatory molecules in the monocytic fraction is aligned with recently reported murine data in which T cell activation occurred in immune competent mice treated with RV alone or RV combined with a PI *(22)*. Because polarized monocytes, in addition to inducing T cell responses, can also act as scavenger cells, we tested whether, besides virus delivery to MM cells, CFZ could also potentiate RV-infected monocytes to phagocytize cancer cells. For this purpose, human PBMCs from 3 different healthy donors (HDs) were treated with RV and CFZ alone or in combination. After an overnight incubation, CD14+ populations from each treatment group were purified and co-cultured with Gfp+ MM cells (MM1S) for 24hrs. Flow cytometry–based analysis showed increased phagocytotic activity when CD14+ cells were pre-treated with RV, as shown by the surge of a double positive (CD14+/Gfp+) cellular population **(Fig. 3L, M)**. A further increase in the double positive population was observed when monocytes were co-treated with RV+CFZ versus RV alone (p=0.028). Significantly higher MM cell death was also observed when the CD14+ fraction was pre-treated with RV+CFZ compared to each single agent using the same experimental settings **(Fig. 3N)**. Aligned with our previous observation, CD14+ cells treated with RV+CFZ showed higher viral replication compared to levels in those treated with RV alone (n=3 donors p=0.006) **(Fig. 3O)**.

### RV combined with CFZ increases viral replication in the bone marrow of MM patients

We previously showed in a phase 1 trial that viral genome is found in the MM cells of relapsed patients treated with single agent Pelareorep, but neither active RV replication nor significant clinical response *(17)* was observed. Because our preclinical data showed that PI increased infection of monocytes and subsequent delivery of virus to the MM cells, we tested this concept in specimens obtained as part of a phase 1b study of RV in combination with CFZ in relapsed myeloma (NCT02101944). Patients were treated on days 1, 2, 8, 9, 15 and 16 of a 28-day cycle. Pre-treatment samples (baseline) were collected just prior to cycle 1 day 1 RV+CFZ infusion for each patient. Treatment included intravenous dexamethasone followed by intravenous CFZ over 30 minutes, and then RV infusion over 60 minutes. We aimed to identify the maximum tolerated dose of Pelareorep combined with CFZ. All safety analyses were conducted during cycle 1. Patients were infused with CFZ 20 mg/m^2^ on days 1 and 2 of cycle 1, and 27 mg/m^2^ thereafter. The starting dose of Pelareorep was 3 × 10^10^ TCID_50_/day, and all patients received dexamethasone 20 mg on each treatment day **(Fig. 4A)**.

**Figure 4.**
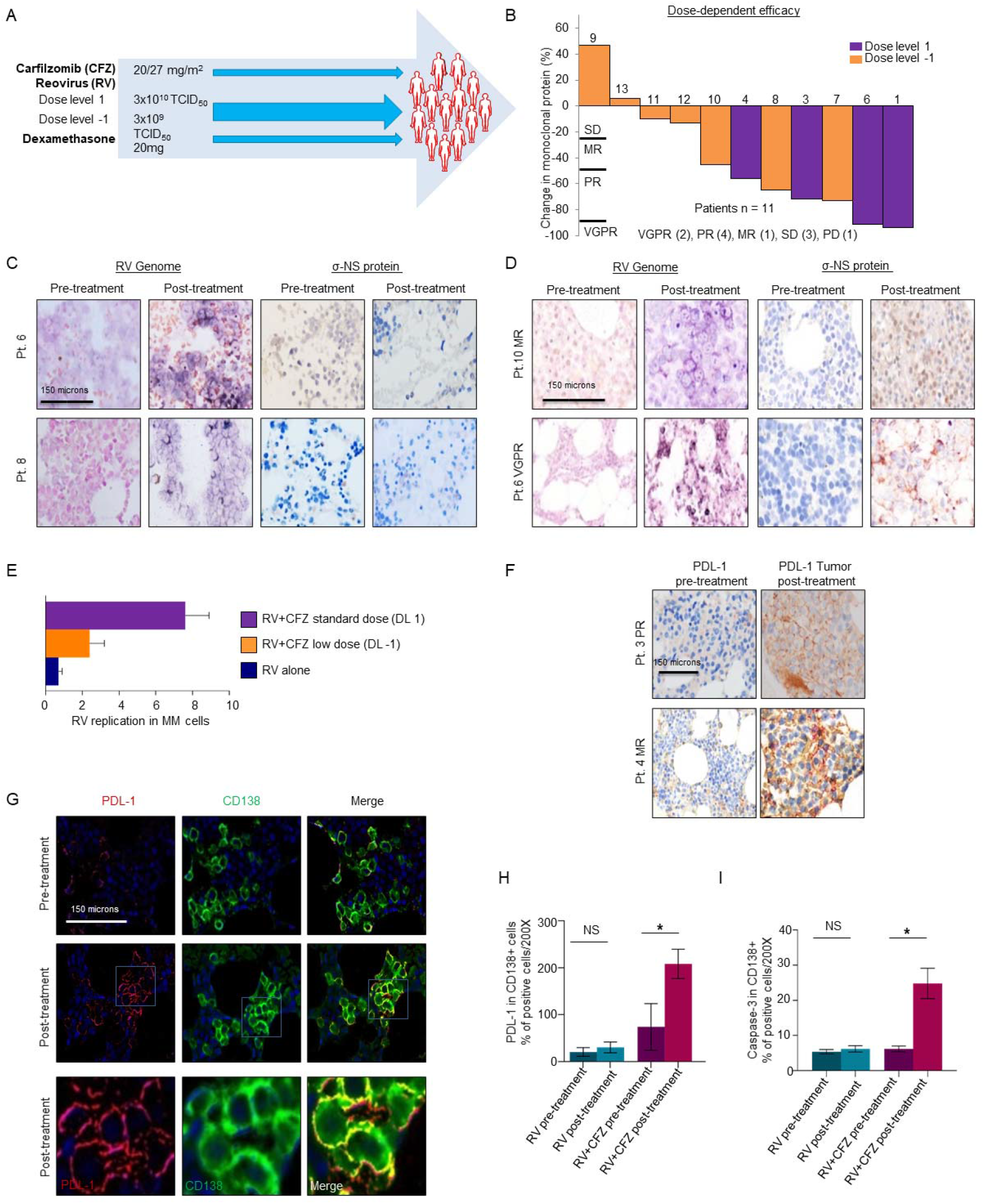
RV combined with CFZ increases viral replication in the bone marrow of MM patients. **(A)** Schematic representation of the treatment schedule for patients with relapsed MM enrolled in Phase 1b clinical trial of the combination of Pelareorep plus carfilzomib; (**B)** Waterfall plot illustrating best response of each patient. The overall response rate (ORR) was 53.8% (7/13) and clinical benefit rate (CBR) was 69.2% (9/13). Patients treated at dose level 1 had an overall response rate (ORR) of 83.3% (5/6) and clinical benefit rate (CBR) of 100%. Responses at this dose level included very good partial response (VGPR, n = 2), partial response (PR, n = 4), minimal response (MR, n = 1) stable disease (SD n=3), and progressive disease (PD n=1). Patients treated at dose level -1 had an ORR of 28.6% (2/7) and CBR of 42.9% (3/7); (**C-D)** 4X magnification images of immunohistochemistry (IHC) showing the in situ data for the detection of reoviral RNA (signal blue with pink counterstain) and reoviral capsid protein (signal brown with blue counterstain) pre- and post-treatment in RV alone and RV+CFZ treated patients. Note that reoviral RNA is evident only after post treatment and that many more cells have detectable viral RNA compared to the capsid protein; (**E)** Bar graph showing σ-NS protein detection at a low and standard dose of CFZ (p=0.028) in the BM. Each value represents the number of positive cells per 200x field; (**F)** 4X magnification images of IHC of PD-L1 protein (signal brown with blue counterstain) pre- and post-treatment (n=5); (**G)** 4X magnification images of immunofluorescence showing the co-expression of PDL-1 (fluorescent red) and CD138 (fluorescent green), and 20X magnification at post-treatment Merged image with co-expression seen as fluorescent yellow (scale bars at 150 micrometers); (**H-I)** Bar graph showing significant upregulation of PD-L1 (H) and Caspase-3 on the surface of MM cells (I) (p=0.005) in RV+CFZ treated patients relative to pre-treatment (n=5) an effect was not observed in the RV-only treated patients. Each value represents the number of positive cells per 200x field.

Thirteen patients were enrolled; baseline demographics are summarized in **Supplementary Table 2**. Seven were male, six were female, and the median age was 60 (range 43 – 70). The median International Staging System stage at diagnosis was 2 (range 1 – 3), and one patient was dialysis dependent. Six patients had evidence of high-risk cytogenetics (+1q21, t(4;14), t(14;16) or del17p) at the time of diagnosis, and ten had high-risk cytogenetic features at the time of screening. The median number of prior therapies was 4 (range 2 – 12), and prior lines of treatment was 2.5 (range 1 – 9). All patients were lenalidomide refractory and BTZ exposed, 84.6% (11/13) were BTZ refractory, two patients were pomalidomide refractory, and one patient was CD38 antibody refractory. Five patients were CFZ exposed, and all were considered to be refractory **(Supp. Table 3)**; 3 of these patients had evidence of disease progression during CFZ treatment, while two others were deemed carfilzomib refractory based on lack of achieving response while on treatment *(28)*. All patients previously CFZ exposed were treated per historical standard of care dosing including treatment on days 1, 2, 8, 9, 15, and 16 of 28-day cycles (20 mg/m^2^ on cycle 1 days 1 and 2, followed by 27 mg/m^2^ thereafter), and three patients had received two prior CFZ-containing regimens. The median duration of exposure to CFZ in these five patients was 8 months (range 1.5 – 20), and the best overall response to a CFZ-containing regimen in these patients was a partial response.

The ten most common treatment-emergent toxicities per CTCAE v5.0 in cycle 1 included hypertension (one grade 2 and four grade 1), thrombocytopenia (two grade 3, one grade 2, one grade 1), anemia (one grade 2 and 3 grade 1), dyspnea on exertion (one grade 2, three grade 1), myalgia (three grade 1), fever (one grade 2, one grade 1), lymphopenia (one grade 3 and one grade 1), nausea (one grade 2, one grade 1), and diarrhea (one grade 1, one grade 2) **(Supp. Fig. 4A)**. Two patients (ID 2 and ID 5) experienced dose limiting toxicities, specifically thrombocytopenia (with bleeding) and acute congestive heart failure, definitely attributable to carfilzomib and possibly related to Pelareorep, respectively. Because of these toxicities, subsequent patients were treated at dose level -1 with CFZ 20 mg/m^2^ and Pelareorep 3×10^9^ TCID_50_/day on all treatment days.

Eleven patients completed at least one cycle of treatment and were evaluable for response. The response outcomes from baseline include very good partial response (VGPR, n=2), partial response (PR, n=4), minor response (MR, n=1), stable disease (SD, n=3), and progressive disease in one patient **(Fig. 4B, and Supp. Fig. 4B)**. The two patients that experienced a DLT following two doses of combination treatment had a 96% (patient ID 2) and 27% (patient ID 5) reduction in measurable disease.

In those patients with BTZ-refractory disease, responses included VGPR (n=2), PR (n=4), and SD (n=3). Responses in patients previously treated with CFZ included PR (n=1), MR (n=1), and SD (n=3). Including all patients evaluable for response, those treated at dose level 1 (DL1) had deeper responses than those treated at dose level -1 and remained on treatment for a longer period of time (mean days of treatment 331.6 [range 84-576] vs 54.5 [range 28-84], respectively), findings that suggest a dose-dependent effect **(Fig. 4B)**. BM aspirates of each patient enrolled in both single agent RV and CFZ+RV trials were collected at baseline and after a week of treatment to assess RV infection (viral genome) and replication (viral capsid) in the BM of treated patients. While active RV replication was not observed in our trial of single-agent RV, as we previously reported *(17)*, as clearly shown by the presence of the viral genome but not capsid protein **(Fig. 4C and Supp. Fig. 4C)**, in the CFZ+RV treated patients, both RV genome and capsid were found **(Fig. 4D and Supp. Fig. 4D)**. Our data showed increased capsid expression in the BM of patients treated with a low dose CFZ (DL-1) compared to baseline (p=0.004), and this effect was further potentiated in patients treated with a standard dose of CFZ (DL1) (p=0.028) **(Fig. 4E)**. Consistent with the preclinical data of Kelly et al. *(21)*, we also found a significant upregulation of the checkpoint inhibitor PD-L1 in five post treatment BM biopsies obtained from patients treated with RV+CFZ (p=0.005), compared to the baseline at pre-treatment **(Fig. 4F, G, H and Supp. Fig. 4E, F)**. This upregulation was not observed in the biopsies obtained from our trial of RV monotherapy **(Fig. 4H)**. Significant caspase-3 activation was also found in tumor biopsies of patients treated with RV+CFZ, compared to the baseline levels (p=0.005, n=5) **(Fig. 4I)**. Caspase-3 activation was not found in the available longitudinal biopsies of patients treated with RV alone (n=5), further supporting active viral replication and subsequent MM cell killing.

### RV combined with PI activates monocytes, expands T cells, and suppresses regulatory T cells

To assess whether RV combined with PI treatment could induce changes in the monocytic and T cell compartments of patients with MM, as we observed in *ex vivo* studies, we performed longitudinal flow cytometry analysis of the peripheral blood (PB) obtained from RV+CFZ treated patients collected at pre-treatment (immediately before C1D1) and during the course of treatment (C1D2-9-15-22-28; C2D1; C3D1). In the first week of treatment, we observed a significant increase in the number of overall monocytes (p=0.006) **(Fig. 5A, B)**, an increase that was mainly due to the expansion of the Classical phagocytic monocyte compartment (CD14++CD16neg) (p=0.03) **(Fig. 5C)**. Aligned with our preclinical data, a significant increase in activated monocytes (CD69+) was also observed (p=0.047) **(Fig. 5D, Supp. Fig. 5A)**. Analysis of the T cell compartment revealed a strong increase in CD8+ T cell frequency upon treatment, especially up to C1D9 (p=0.04) **(Fig. 5E and Supp. Fig. 5B)** and correlated with a substantial reduction of CD4+ cells (p=0.03) **(Fig. 5F and Supp. 5C)**, as well as a significant reduction upon treatment of the CD4/CD8 ratio (p=0.04) **(Fig. 5G**, see also below).

**Figure 5.**
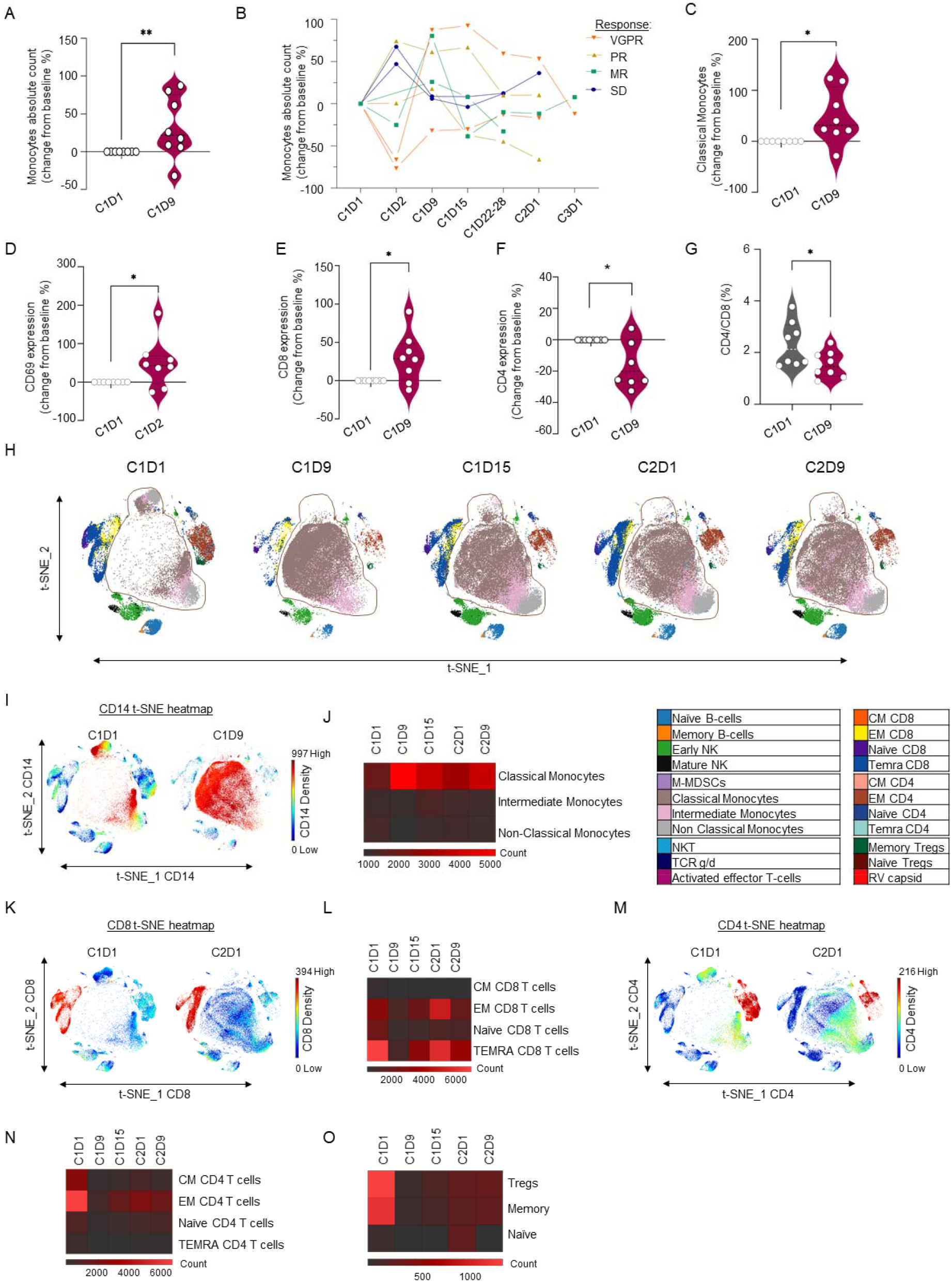
RV combined with a PI activates monocytes, expands T cells, and suppresses regulatory T cells. **(A)** Violin plots representing multiparametric flow cytometry studies on PB from MM relapsing patients enrolled in RV+CFZ Phase 1b clinical trial, showing overall expansion of total monocytes absolute count (**p≤0.01); (**B)** Line graphs representing longitudinal multiparametric flow cytometry studies on PB from the trial, showing overall expansion of total monocytes absolute count. Wilcoxon signed rank p-values: C1D9 > C1D1 p-value= 0.054; (**C-D)** Violin plots representing higher frequency of CD14++CD16neg Classical Monocytes (C) and increase in CD69 activation marker in the total monocytes (p=0.047) (D) on treatment up to the C1D9 compared to baseline C1D1. Statistical analysis was performed following Wilcoxon signed rank p-values, **p≤0.01, *p≤0.05; (**E-F)** Violin plots highlighting the increased CD8 expression (E) and the decreased CD4 expression (F) up to C1D9. Data are expressed as change from baseline %, Wilcoxon signed rank p-values: *p≤0.05; (**G)** Violin plot highlighting decrease in CD4/CD8 ratio from baseline up to the C1D9. Data are expressed as change from baseline %, Wilcoxon signed rank p-values: *p≤0.05; **(H)** In-depth immune profiling of a longitudinal CFZ-resistant patient enrolled in RV+CFZ Phase 1b was performed with the Maxpar Direct Immune Profiling System using a dry 30-marker antibody panel. Hierarchical clustering and statistical mapping performed algorithmically via the Cytobank© platform. vi-SNE analysis (iterations=1000, perplexity=30) displayed in 2D plots using the resultant t-SNE 1 and t-SNE2 dimensions. High-fidelity FlowSOM (“self-organizing map”) (metacluster=10 and cluster=100) based on vi-SNE 2D plots showing 22 immune different immune-compartments; (**I-J)** t-SNE heatmap highlighting expression of selected monocyte population after treatment C1D9 (I) and heatmap showing absolute count of the different monocyte compartments (J); (**K-L-M-N)** t-SNE heatmaps highlighting expression of selected CD8 (K) and CD4 (M) populations comparing the pre-treatment C1D1 with the post-treatment C2D1, and heatmaps showing the different CD8 (L) and CD4 (N) immune subsets longitudinally; **(O)** Heatmap showing overall distribution of naïve and memory Tregs during the course of the therapy.

In line with our multiparametric flow cytometry studies, CyTOF analysis in a CFZ-resistant patient indicated clear monocyte and T cell expansion, as shown by cell cluster visualization of the different immune populations **(Fig. 5H)**. Specifically, high-dimensional t-SNE heatmaps and FlowSOM revealed a dramatic increase in the CD14+ monocytic fraction **(Fig. 5I)**. Subcluster characterization indicates a specific increase in the classical phagocytic monocytes **(Fig. 5J)**, with the major peak after one week of treatment (from C1D1 [total cell count 13,385] to C1D9 [total cell count 53,835]) **(Fig. 5I, J and Supp. Fig. 5D)**. As expected, the increase in activated monocytes was followed by a robust increase of CD8+ cytotoxic T cells and a decrease in total CD4+ T cell numbers. Specifically, an increase in CD8+ EM T cells up to C2D1 (C2D1 count: 4964 versus pre-treatment C1D1 count: 2745) **(Fig. 5K, L and Supp. Fig. 5E)** coincided with a strong decrease in CD4+ T cells (from C1D1 count: 14893 to C2D9 count: 4523) **(Fig. 5M, N and Supp. Fig. 5E)**. Circulating regulatory T cells (Tregs) were sharply reduced after the first cycle of therapy (from C1D1 count: 1405 to C2D1 count: 293). Moreover, an in-depth classification of this population showed the same trend in both memory and naïve Tregs **(Fig. 5O and Supp. Fig. 5F)**. Collectively, our data indicate a robust subset-specific phagocytic monocyte profile in the circulation of Pelareorep+CFZ□treated patients, followed by a cytotoxic CD8+ T cell expansion.

### RV combined with CFZ treatment promotes T cell diversity in MM patients

Because we observed an increase in cytotoxic CD8+ T cells which followed a monocytic expansion, we then investigated whether T cell proliferation induced by RV+CFZ could also be associated with changes in T cell clonality. Therefore, we analyzed the T cell receptor (TCR) repertoire in the blood of RV+CFZLJtreated patients when material was available for the analysis (n=9) within the first two weeks of treatment *(29, 30)*. All samples were immune-sequenced on a multiplex PCR high-throughput TCR sequencing assay at deep resolution **(Supp. Fig. 6A)**. Specifically, we sequenced the variable (TCRβV) and joining (TCRβJ) regions in the complementarity-determining region 3 (CDR3) of peripheral blood T cells.

Although we did not observe significant diversity at baseline **(Supp. Fig. 6B, C)**, 78% of the subjects (7 of 9) showed an increase in peripheral clonality with treatment **(Fig. 6A)**. Using a linear mixed effects model to control for variation in baseline clonality, we observed a trend toward an increase in overall clonality with treatment (p=0.07) and lower overall clonality in those patients with an objective response (p=0.06), reflecting less monoclonal expansion in response to therapy **(Fig. 6A)**. Moreover, 7 out of 9 subjects showed a decrease in peripheral richness with treatment (p=0.04) **(Fig. 6B)**, an effect that was consistent with previous Pelareorep studies *(31)*.

**Figure 6.**
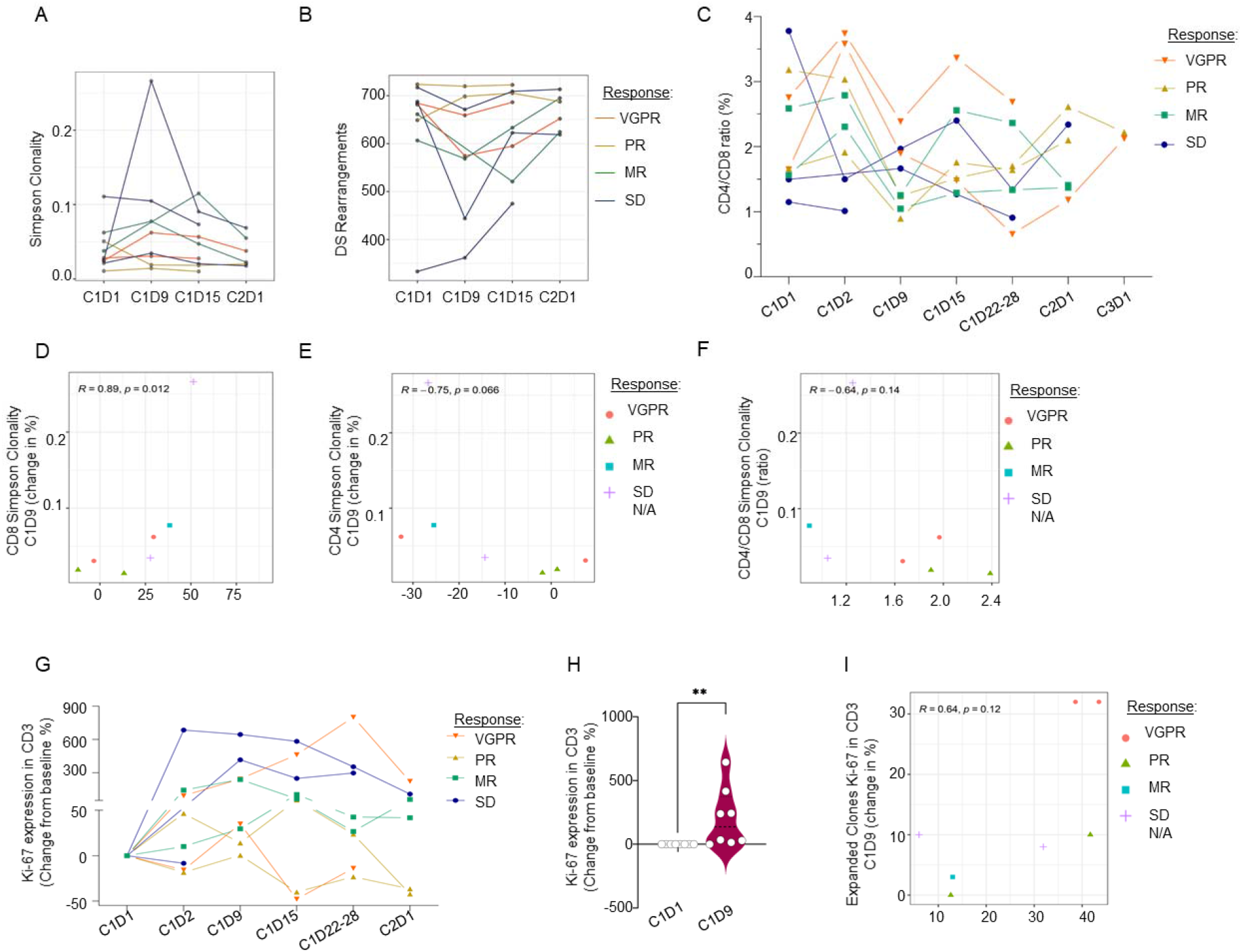
RV combined with CFZ treatment promotes T cell diversity in MM patients. **(A-B)** Analysis of the peripheral TCR repertoire from baseline (C1D1) and on-treatment (C1D9, C1D15, and C2D1) calculated for the clone’s distribution. Simpson clonality showing in 7/9 subjects an increase in peripheral clonality at C1D9 or C1D15 on treatment relative to C1D1 (A), and line graph describing Peripheral Richness Changes calculated as the number of unique rearrangements after down-sampling to a common number of T cells (n=735), showing in 7/9 subjects a decrease in peripheral richness at C1D9 or C1D15 on treatment relative to C1D1 (B); (**C)** Line graph representing longitudinal multiparametric flow cytometry studies on PB from relapsed MM patients enrolled in RV+CFZ Phase 1b clinical trial showing CD4/CD8 ratio and Wilcoxon signed rank p-values: C1D9 < C1D1 p-value = 0.04; C1D22-28 < C1D1 p-value = 0.004; C2D1 >C1D1 p-value = 0.05. (**D)** Scatterplot showing correlations between the clonality metric with the CD8 expression from FACS analysis (a lower increase in CD8 is associated with lower clonality and responders at C1D9). **(E)** Scatterplot showing correlations between the clonality metric with the CD4 expression from FACS analysis (less of a decrease in CD4 is associated with lower clonality and responders at C1D9); **(F)** Scatterplot showing correlations between the clonality metric with the CD4/CD8 expression from FACS analysis, suggesting that a high CD4/CD8 ratio may be associated with better response post-treatment; **(G-H)** Line graph representing longitudinal multiparametric flow cytometry studies on the same set of patients showing Ki-67 expression in total T cells after treatment (G) and violin plot highlighting the increased Ki-67 expression up to the C1D9 (H). Data are expressed as change from baseline %, Wilcoxon signed rank p-values: C1D9> C1D1 p-value = 0.004; C1D15 >C1D1 p-value = 0.07; C1D22-28 >C1D1 p-value = 0.07; (**I)** Scatterplot showing correlations between the expanded clonality with the Ki-67 measures in total CD3 compartment from FACS analysis showing that subjects with high Ki67 have more clonal expansion.

Since recent studies have revealed that the TCR repertoire is largely different between CD8+ and CD4+ T cells *(32)*, we also performed an in-depth comparison between multiparametric flow cytometry analysis of the CD4, CD8 and Ki67 measures and immunoSEQ metrics (clonality, richness, T cell fraction and clonal expansion).

As lower CD8 clonality is often associated with better outcomes, we observed that the two patients experiencing VGPR showed a higher CD4/CD8 ratio associated with lower CD8 clonality at C1D9, **(Fig. 6C, D, E, F)**, suggesting that a better response may depend on both CD4+ and CD8+ T cell responses **(Fig. 6D, E, F)**. Of particular importance, we compared TCR measures with our flow cytometry data, which exhibited an increased expression of the cell cycle marker Ki-67 in blood lymphocyte T cells on treatment (C1D9), compared to baseline levels (C1D1) (p=0.004) **(Fig. 6G, H, I and Supp. Fig. 6D, E)** and was associated with a strong orthogonal metrics correlation with peripheral clonal expansion, especially in the two subjects with VGPR **(Fig. 6G)**. Aligned with these data, immunohistochemistry of BM core biopsies showed a trend towards CD8+ T cell recruitment on treatment (8.5% to 26.8% positive cells per high power field, p=0.060) **(Supp. Fig. 6F, G)**. To assess whether the addition of CFZ may contribute to support T cell specific responses against RV-infected cancer cells, we performed an ELISpot assay to measure IFN-γ production after stimulation of mouse splenocytes isolated from mice treated with RV and CFZ alone or in combination. Specifically, 2×10^6^ 5TGM1 cells were intravenously injected in syngeneic C57BL/KaLwRij mice. After 9 days from the injection, mice were randomly divided in four treatment groups. Mice were treated once a week with intravenous injection of RV alone (2×10^7^ PFU) (n=5), biweekly with intraperitoneal injection of CFZ (1.6 mg/kg) (n=6), or the combination of both (n=6) (**Supp. Fig. 6H**). Diluent (PBS 1X) treatment of mice was used as control (n=8). Our data show that mice treated with RV+CFZ have a higher percentage of circulating effector memory (EM) CD8+ T cells when compared to the other treatment groups **(Supp. Fig. 6I, J, K)**. After 3 weeks, mice were humanely sacrificed, and IFN-γ production was measured in isolated splenocytes stimulated with RV-infected and non-infected 5-TGM1 cells. Our data show that animals treated with RV+CFZ have significantly higher production of IFN-γ when stimulated with RV-infected cells (**Supp. Fig. 6L, M**), an effect that was not found in mice treated with the single agents alone (**Supp. Fig. 6N**). These data further support that the addition of CFZ plays a pivotal role in orchestrating T cell responses against viral infected MM cells and in the anti-MM response.

## DISCUSSION

Here we report that RV actively infects and replicates in JAM-1(+) circulating monocytes, which in turn delivers active replicative virus to MM cells. Our data provide clear evidence that the anti-viral inflammatory signals in monocytes primarily rely on the NF-kB activation pathway, which is almost completely curtailed by the addition of PIs. We also show that the addition of a PI strongly improves RV infection and replication in canonical phagocytic monocytes (see **Fig. 7** for a mechanistic illustration). These data are aligned with reports showing the importance of NF-kB activation in monocytes after RNA virus infection, including SARS-CoV-2 and respiratory syncytial virus *(33, 34)* and are consistent with the well-known anti-MM activity of PIs, which rely on blocking aberrant NF-kB signals in MM cells *(35) (36)*.

**Figure 7.**
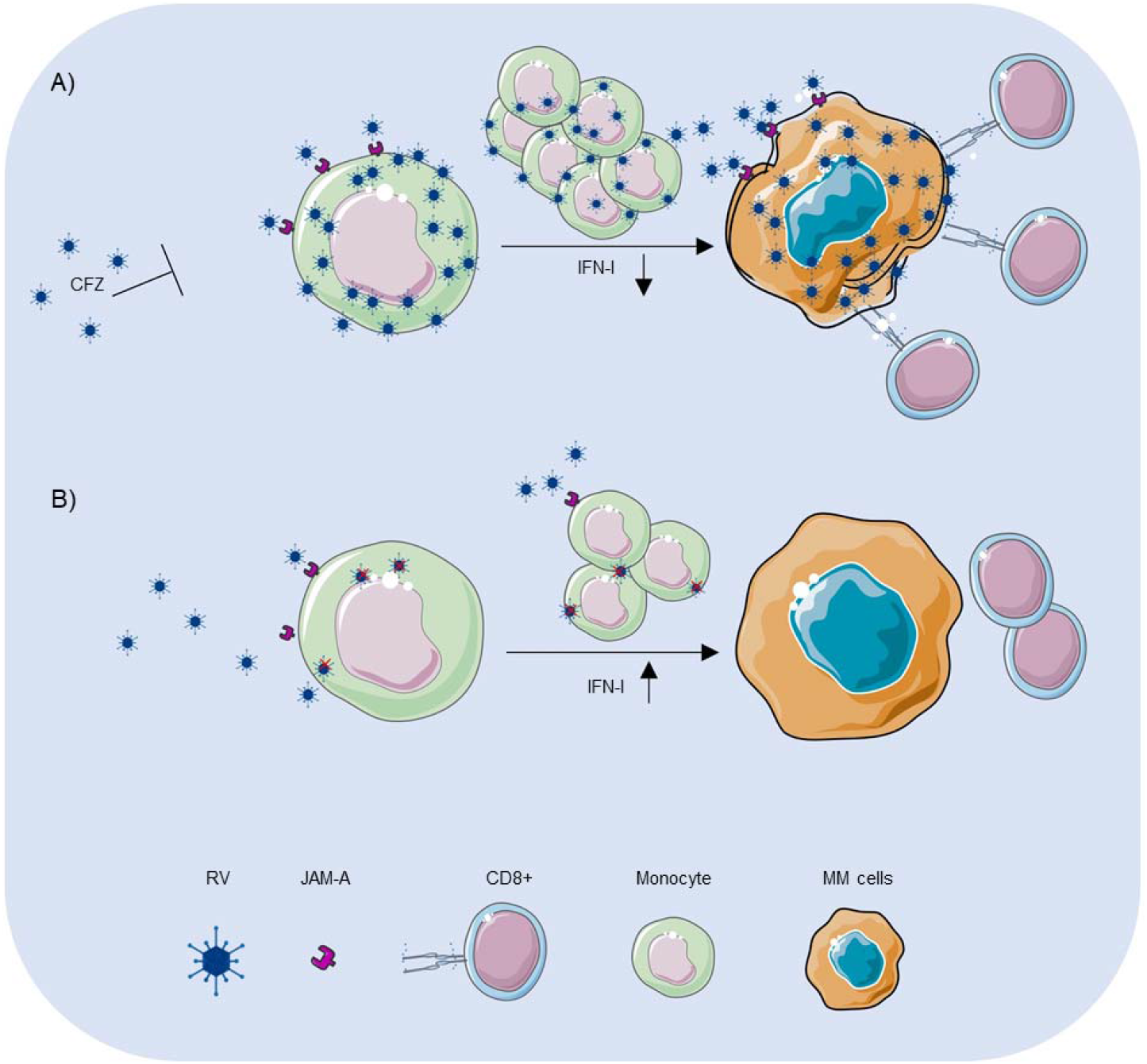
Graphical abstract illustrating the proposed mechanism of RV combined with CFZ in patients with MM. **(A)** Graphical representation of the proposed mechanism showing how proteasome inhibition (CFZ) increases the viral delivery to MM cells and enhances myeloma oncolytic reovirus therapy by suppressing IFN-I monocytic anti-viral immune responses through monocyte expansion and CD8+ cytotoxic T cell activation against cancer cells; **(B)** Graphical representation of the proposed mechanism of the lack of viral delivery to MM cells in the absence of proteasome inhibition.

Although PIs have been successfully combined with both immunomodulatory and antibody-based therapies in MM *(37)*, their characterization as immune suppressive drugs has excluded their combination with novel T cell-based therapies. PIs are thought to reduce T cell activity based on preclinical (36) and clinical data showing an increase in the risk for varicella zoster reactivation *(38)*; the molecular mechanism behind this observation has not yet been characterized.

Here we show that, both preclinically and clinically, that blunting the anti-viral inflammatory response with a PI did not impair monocytic activation nor T cell activation and expansion upon RV infection, and even increased phagocytic activity of the monocytic fraction against RV-infected MM cells. Hence, in the context of oncolytic viruses, in which delivery to the tumor site is one of the main roadblocks to fully translate their potential anti-cancer benefits into the clinic, the addition of a PI is a logical next step to impair monocytic anti-viral responses and increase viral delivery to the cancer cells, in which higher viral replication and cytolytic activity can be achieved.

Further clinical benefits of adding PIs to viral oncolytic therapies may also derive from their ability not only to potentiate RV infection of cancer cells but also to support RV-induced immune activation in immunosuppressed patients. The dual immune modulatory effect of PIs may be explained by one or both of the following: a) the T cell response is mainly driven by the JAK-STAT pathway *(39, 40)*; b) PI treatment does not affect IFN-γ release upon viral infection. Of note, although our data seem to contradict recently published data showing that PI induces anti-viral IFN-I signaling in MM cells *(41)*, mimicking a viral attack, we observed that in human circulating monocytes PI treatment alone induced transcriptional activation of IFN-α and IFN-β (**Supp. Fig. 3C, D**), but that this induction was significantly lower than what was observed upon RV infection. IFN-I activation signaling upon PI treatment was detected only in the absence of activated NF-kB pathways by exogenous signals, supporting that PIs can either enhance or impair IFN-I signaling through different signaling pathways. Aligned with Gulla et al. *(41)*, we found that PIs do not impair monocytic or T cell activation, positioning it as an ideal companion drug in the setting of an oncolytic virus or even a T-cell directed therapy such as chimeric antigen directed T-cells or bispecific antibodies targeted to CD3. In fact, the goal in oncolytic virus therapies in both hematological malignancies and solid tumors is to allow maximal infection of tumor cells, and activation of the patient’s immune system to clear them once they are infected.

In MM, oncolytic viral therapies are still limited; in fact, despite promising preclinical results with myxoma, varicella, and adenovirus, only an engineered measles virus and reovirus have been given to patients with relapsed MM.

Concerns about intravenous RV injection, or any oncolytic virus, in hematologic malignancies often stem from the abrupt production of antiviral antibodies that are anticipated to neutralize intravenous virus. These common concerns are not supported by several published data showing that immune cells (T cells or dendritic cells) can be loaded with RV *ex vivo* and administered systemically to deliver virus to tumors, even in the presence of anti-reovirus neutralizing antibodies *(42, 43)*. Aligned with these data, results in patients with colorectal liver metastases indicated that free RV delivered systemically could access tumors, and that functional virus was associated with immune cells in the blood but was not found in plasma *(23)*. Neutralizing antibodies can instead be used by RV to internalize in human CD11b+ monocytes, which later can deliver replicative RV to tumor cells, resulting in infection and ultimately lysis *(44)*. Other published data in an animal model have clearly shown that increasing the frequency of circulating monocytes through GM-CSF treatment enhances delivery and activity of intravenous RV *(45)*, further supporting that enhancing RV infection of circulating monocytes independently of the presence of neutralizing antibodies may be a successful therapeutic strategy.

Another concern is that non-engineered naturally occurring viruses such as RV are unlikely to be therapeutically active unless partnered with an activating agent. PIs are a rational choice to combine with viruses, as they prevent signaling through nuclear factor NF-κB in immune cells, abrogating the initial inflammatory antiviral response, even though this effect is short-lived *(46)*. Nevertheless, most studies of PIs in combination with an oncolytic virus focus on induction of the unfolded protein response and its potential enhancement of cytotoxic effects *(47)* or viral replication *in vitro (48)*. The first publication investigating the combination of RV plus the PI BTZ reported that the addition of BTZ enhanced RV-induced apoptosis in MM cells *(21)*. A subsequent publication investigating this combination could not evaluate for synergy, as only doses that led to 50% cytotoxicity were used. However, this study emphasized the importance of RV infection of the tumor microenvironmental cells *(22)*. Our experiments showed that at early time points, PI treatment of MM cells did not enhance viral replication or improve RV-induced apoptosis, but rather showed a decrease in viral replication and non-additive effect in tumor cell killing. This is a result that we believe is consistent with the tendency of viruses to actively replicate and produce viable progeny particles in living host cells rather than in dying cells *(49)*.

Our clinical data clearly show that the addition of a PI plays a pivotal role in supporting active viral replication in the cancer cells of patients with MM. In our patients treated with combination treatment, viral replication was associated with the presence of both viral genome and viral capsid in the BM and caspase-3 activation, an effect that was not observed in our previous single agent RV trial. In line with our *ex vivo* testing, patients with MM who were treated with Pelareorep+CFZ showed monocytic and T cell expansion and activation, an immunological and clinical response that was observed even in patients resistant to CFZ, further highlighting the immunomodulatory activity of a PI independent of the direct anti-MM effect. Although the attribution of a specific clinical response to one agent in a combination therapy can be challenging, our animal data show that addition of a PI is essential to orchestrate T cell responses to RV-infected cancer cells, further supporting the importance of a PI in oncolytic viral therapy. Aligned with Kelly *et al. (50)*, who reported that RV-infected MM cells significantly upregulate the expression of surface PD-L1, we found that the MM cells of Pelareolep+CFZ treated patients have increased PD-L1 expression on their surface compared to patients treated with Pelareorep alone. Although higher PDL-1 expression is a sign of productive MM cell infection, we have to also take into consideration that PD-L1 upregulation can negatively impact direct killing of cancer cells by T cells. We believe that, as suggested from Kelly *et al*., the combination with anti-PD-L1 therapy can further potentiate the anti-MM activity of Pelareolep, which we plan to explore using combination with CFZ.

In conclusion, our data are the first to highlight proteasome inhibition as an optimal therapeutic companion to enhance oncolytic virus therapy independently of its direct anti-cancer activity, leading these observations to be relevant not only for MM but also for oncolytic viruses in solid tumors.

## MATERIALS AND METHODS

### Study design

We present clinical and correlative data from NCI 9603, a phase 1b trial testing carfilzomib and Pelareorep, the proprietary form of reovirus, in patients with relapsed MM. The Ohio State University Cancer Institutional Review Board (Columbus, OH) approved this study, and informed consent was obtained from all enrolled patients (www.clinicaltrials.gov, NCT 02101944). Patients with relapsed and refractory myeloma according to the International Myeloma Working Group (IMWG) diagnostic criteria for symptomatic myeloma were enrolled (21). Patients must have received prior lenalidomide and bortezomib therapy, progressed on or within 60 days of the most recent therapy, and had an Eastern Cooperative Oncology Group (ECOG) performance score ≤ 2 or Karnofsky Performance Status ≥ 60%. Prior autologous and allogeneic transplantations were permitted.

Patients were required to have measurable disease defined as serum monoclonal protein ≥ 500 mg/dL, > 200 mg of monoclonal protein in a 24-hour urine sample, or serum immunoglobulin free light chain ≥ 100 mg/L with an abnormal kappa to lambda free light chain ratio. Adequate organ and marrow function was required. There was no serum creatinine requirement. Exclusion criteria included congestive heart failure with a LVEF < 50% at the time of screening.

The primary objectives were to 1) determine safety and tolerability, and define the maximum tolerated dose of the regimen; and 2) obtain evidence of reovirus entry into myeloma cells. Patients were enrolled per a standard 3+3 dose escalation schedule.

Correlative studies are included from patients with relapsed MM who were treated with Pelareorep alone in a previously published phase 1 study *(17)*.

### Mice

5-TGM1 murine myeloma cells were harvested during the logarithmic growth phase and injected in 12 immune competent C57BL/KaLwRijHsd mice (0.05 mL/mouse containing 1×10^5^ cells in PBS). Cells were injected intrafemorally, and tumor progression was monitored weekly by mandibular bleeding. CD138+ cells were detected by flow cytometry. On day 28, mice were randomized in 2 different treatment groups. One group was treated with a solution of clodronate-liposomes (dichloromethylene diphosphonate-liposome solution) (Encapsula Nanaoscience LLC, liposomal clodronate Cat. #NC0337390) to induce monocyte/macrophage depletion and then injected intravenously with 1×10^7^ TCID_50_ of RV, while the second group received only Encapsome (plain liposomes for control) solution before RV injection. Control group mice treated only with plain liposomes solution or clodronate-liposomes alone were also included. After 48hrs, mice were euthanized and femurs were collected. BM-MNCs were isolated and processed by flow cytometry to assess RV capsid formation and confirm the expected macrophage depletion.

### Mass Cytometry (CyTOF) Staining and Acquisition

A total of 2-4×10^6^ PBMCs obtained either from MM or healthy donors were stained with a customized panel containing 34 metal-conjugated antibodies (32 surface and 2 intracellular staining and Cell ID cisplatin for non-viable cell detection) **(Supplementary Table 4)** according to Fluidigm’s CyTOF protocols for Cell-ID Cisplatin (PRD018 version 5) (Cat. 201064) and Maxpar Cytoplasmic/Secreted Antigen Staining with Fresh Fix (400279 Rev 05). Non-commercial metal-conjugated antibodies were purchased purified from Biolegend **(Supplementary Table 5)** and in-house conjugated according to Fluidigm’s protocol for Maxpar Antibody Labeling (PRD002 Rev 12). PBMCs derived from Reovirus-treated patients (NCI Protocol 9603 IRB 00105298) were stained with Fluidigm’s Maxpar Direct Immune Profiling Assay Cell Staining (PN 400286 B1). Samples were acquired, exported as FCS files, and normalized on Fluidigm’s Helios (Software 7.0.5189).

### CyTOF Analysis

Non-custom panel analysis was analyzed using Maxpar Pathsetter™ software powered by GemStone 2.0.41, Verity Software House, Topsham, Maine. (version 2.0.45). Custom panel FCS files were manually analyzed using FlowJo™ Software (Windows edition, Version 10.6. Becton Dickinson Company; 2019), and the Cytobank© platform (https://www.cytobank.org) (Cytobank, Inc., Mountain View, CA) for gating and tSNE plots, and FlowSOM analyses for T cells and monocytes.

### T-cell receptor variable beta chain sequencing

Immunosequencing of the CDR3 regions of human TCRβ chains was performed using the ImmunoSEQ® Assay (Adaptive Biotechnologies, Seattle, WA). Extracted genomic DNA was amplified in a bias-controlled multiplex PCR, followed by high-throughput sequencing. Sequences were collapsed and filtered in order to identify and quantitate the absolute abundance of each unique TCRβ CDR3 region for further analysis as previously described *(29, 30, 51)*. The fraction of T cells was calculated by normalizing TCR-β template counts to the total amount of DNA usable for TCR sequencing, where the amount of usable DNA was determined by PCR-amplification and sequencing of several reference genes that are expected to be present in all nucleated cells.

### Statistics

For longitudinal analysis on PB and BM of patients to detect significant differences between pre (C1D1) and post treatment (C1D9) measurements, one-sided Wilcoxon signed-rank tests were performed for matched samples. A p-value < 0.05 was considered as statistically significant. Data were analyzed using R software (RStudio, version 1.4). For *in vitro* experiments, data are reported as mean ± SD of three to four experiments. For *ex vivo* studies, blood samples were collected from each HD and were split into the number of treatments under comparison. Appropriate t tests (paired or independent samples; two-sided or one-sided) or nonparametric tests (Mann-Whitney U test, Wilcoxon signed-rank test) were performed to assess significant differences between two (or more) treatments and/or groups. P-value < 0.05 was considered a statistically significant. Animal data were analyzed by ANOVA followed by Tukey’s multiple comparison tests for pairwise comparison.

Further Materials and Methods can be found in Supplementary Materials

## Data Availability

All data produced in the present study are available upon reasonable request to the authors

## List of Supplementary Materials

Tables S1 to S6

Fig. S1 to Fig S6

Supplemental Methods

## Acknowledgments

The authors wish to thank Dr. Domenico Viola and Dr. Francesca Besi for their technical support in our study.

## Author contributions

CCH & DWS wrote the clinical protocol, enrolled the patients, analyzed the clinical data, and wrote portions of the manuscript associated with the clinical data. GN, AC, AP, MC, JS, GM, and AK analyzed the data. FP and AD designed and performed the preclinical experiments, analyzed the data, and wrote the portions of the manuscript associated with the preclinical data. TT, JW and MS performed CyTOF experiments and assisted with analysis. EC, LN, YZ, MM, and HV performed experiments. All the authors have contributed substantially to revising the manuscript for important content and final approval.

