## Supplemental Information for "Proteasome inhibition enhances myeloma oncolytic reovirus therapy by suppressing monocytic anti-viral immune responses"

**Supplemental Tables**

**Supplemental Table 1.** CyTOF gating strategy

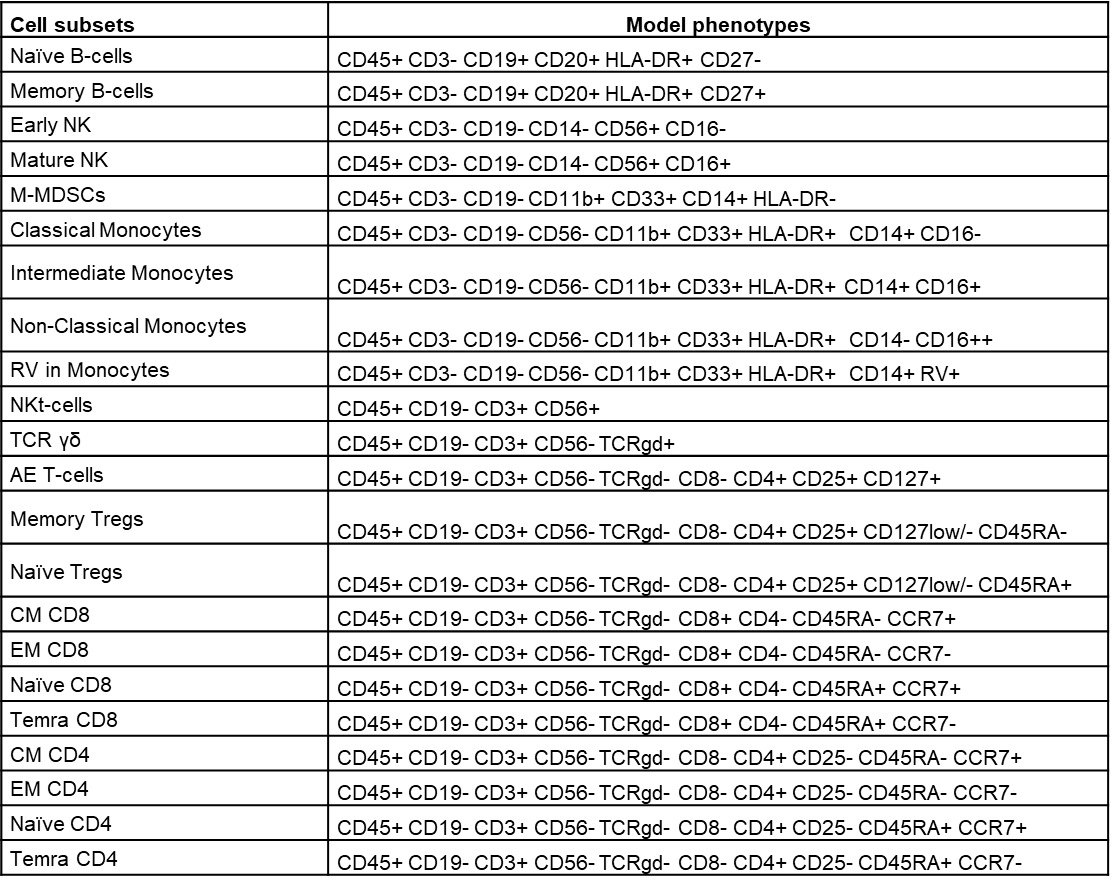

**Supplemental Table 2.** Demographics

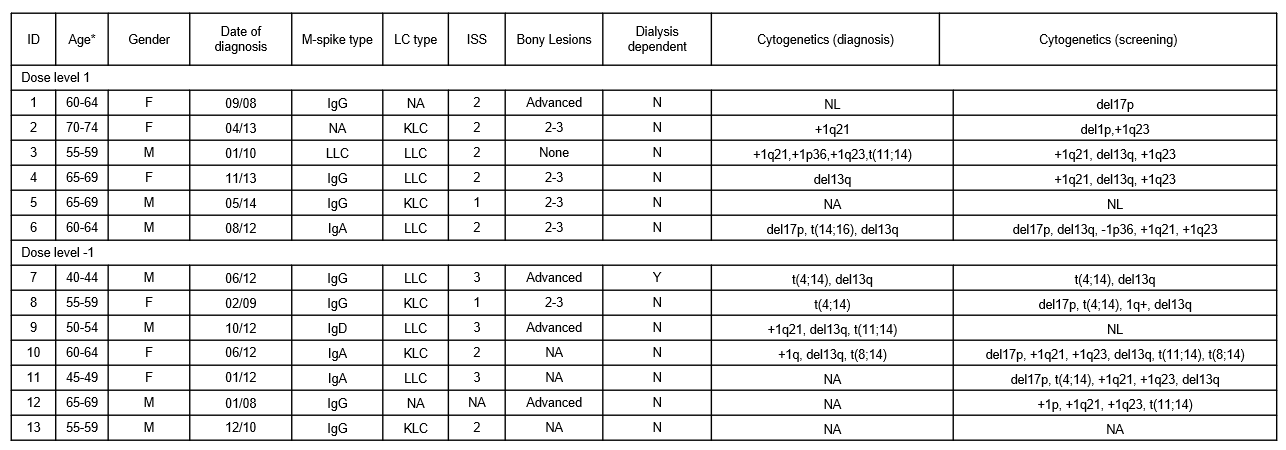

NOTE: Patient demographics of all patients enrolled on the phase 1b Reolysin and Carfilzomib trial. Patient factors included age, gender, and race; date of diagnosis, monoclonal protein and serum light chain type, ISS stage at diagnosis, presence or absence of bony lesions at diagnosis, dialysis dependence, and cytogenetics at diagnosis and screening. Abbreviations: ID = identification, M-spike = monoclonal protein, LC = light chain, ISS = international staging system, F = female, M = male, IgG = immunoglobulin G, NA = not applicable or available, LLC = lambda light chain, KLC = kappa light chain, IgA = immunoglobulin A, IgD = immunoglobulin D, N = no, Y = yes, NL = normal, del = deletion, t = translocation. *Ages are listed within 5-year ranges.

**Supplemental Table 3.** Prior treatments

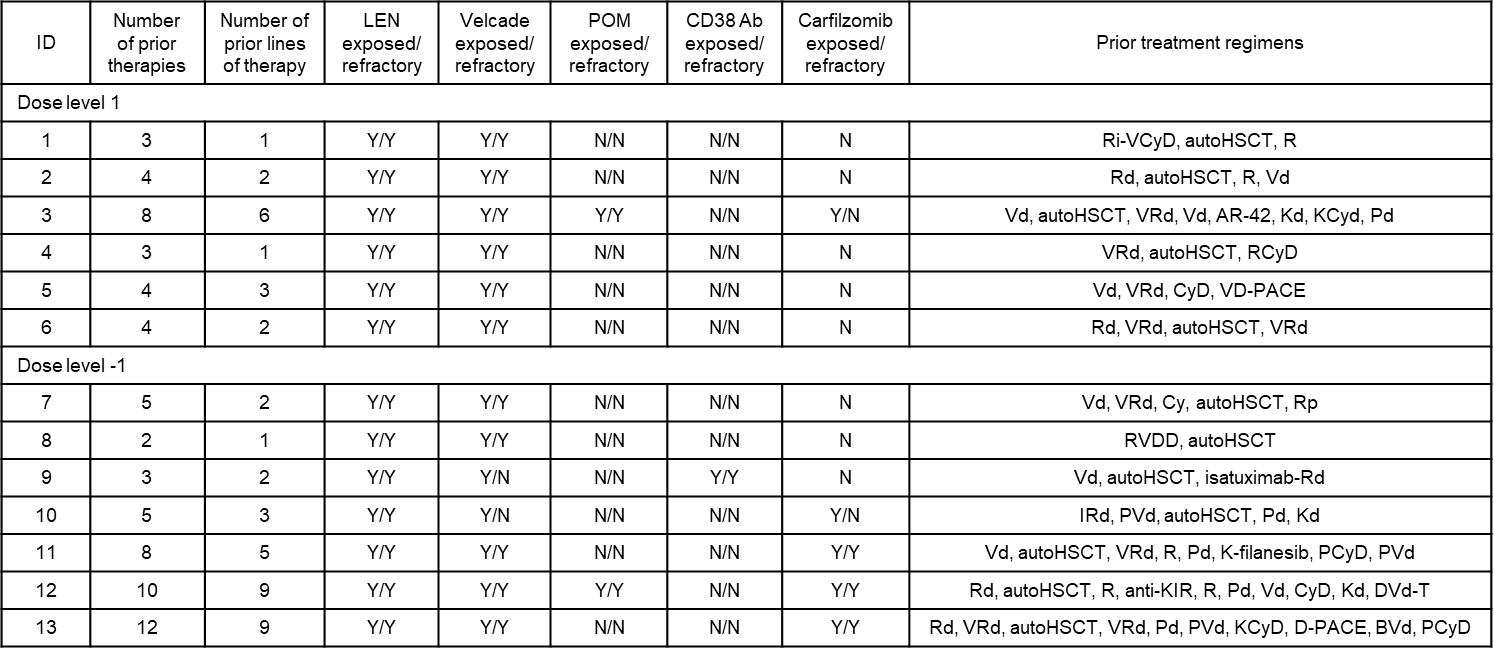

NOTE: Prior treatments and carfilzomib exposure in all patients treated on the phase 1b Reolysin and Carfilzomib trial. Abbreviations: ID = identification, LEN = lenalidomide, POM = pomalidomide, Ab = antibody, Y = yes, N = no, Ri = rituximab, V = velcade, Cy or C = cyclophosphamide, D or d = dexamethasone, autoHSCT = autologous stem cell transplantation, R = revlimid, K = carfilzomib, P = pomalidomide, PACE = cisplatin, doxorubicin, cyclophosphamide, etoposide, p = prednisone, I = ixazomib, T = thalidomide, B = bendamustin

**Supplemental Table 4.** Maxpar direct immune profiling assay 30-marker panel with clones and heavy metals (Fluidigm)

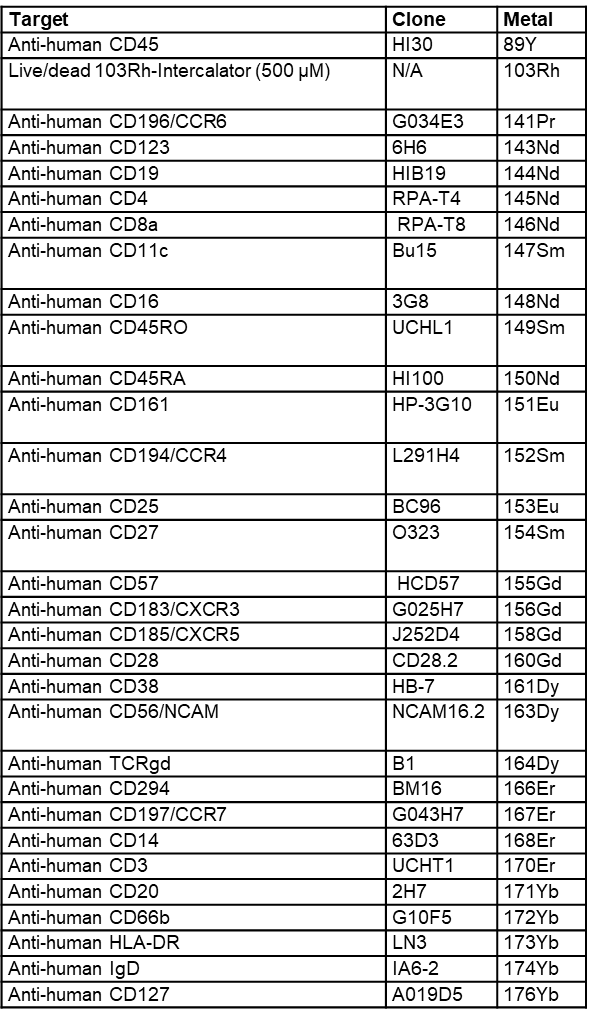

**Supplemental Table 5.** CyTOF customized 36-marker panel

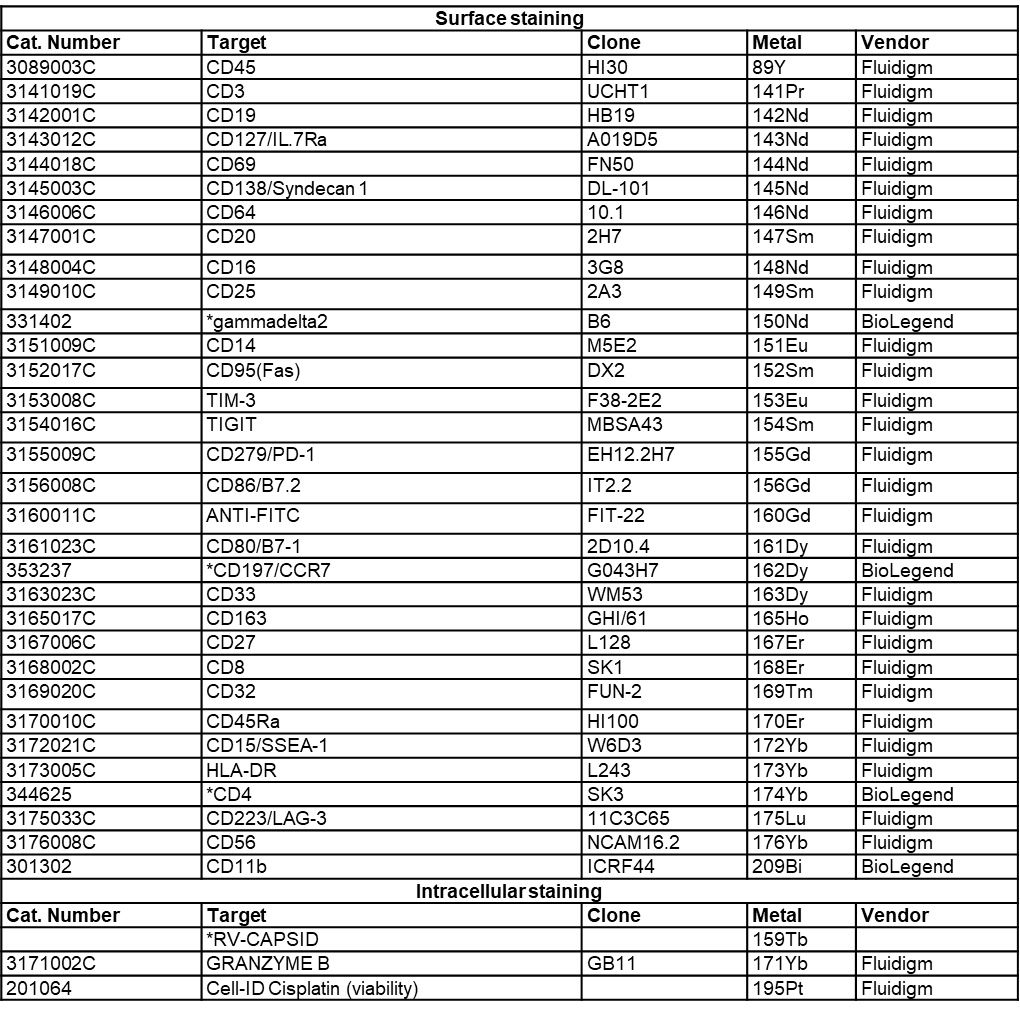

*In-house conjugated antibodies

**Supplemental Table 6.** Flow cytometry surface markers

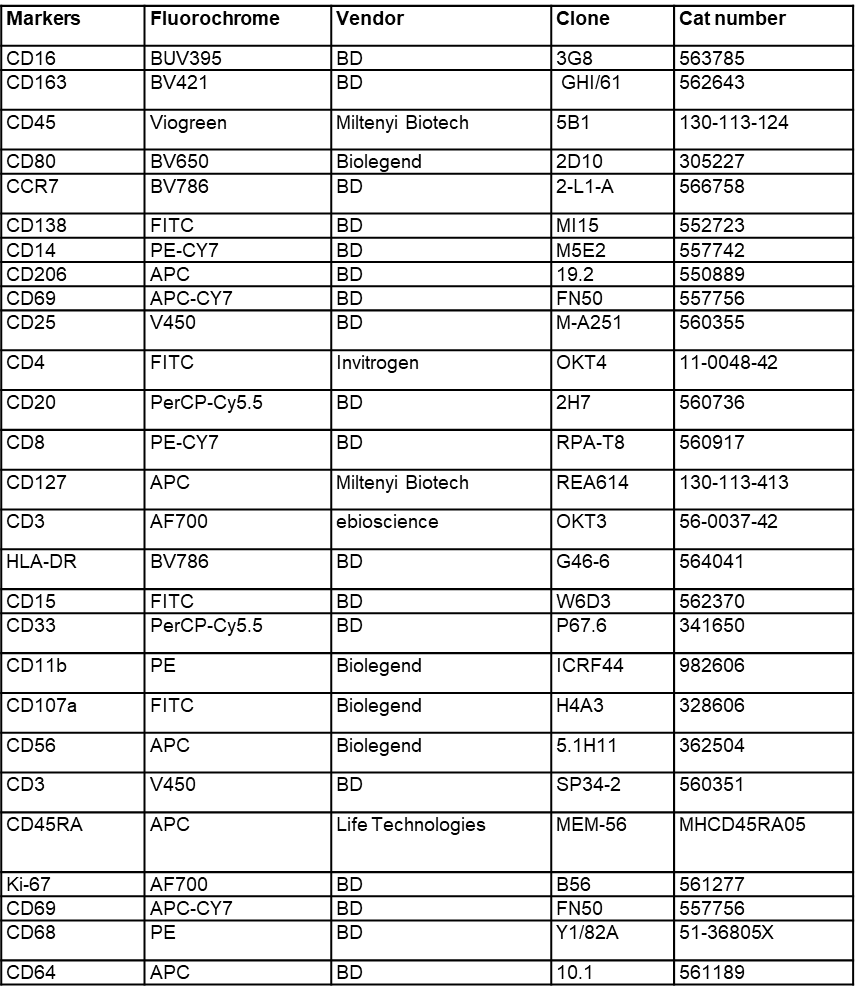

**Supplemental Figures**

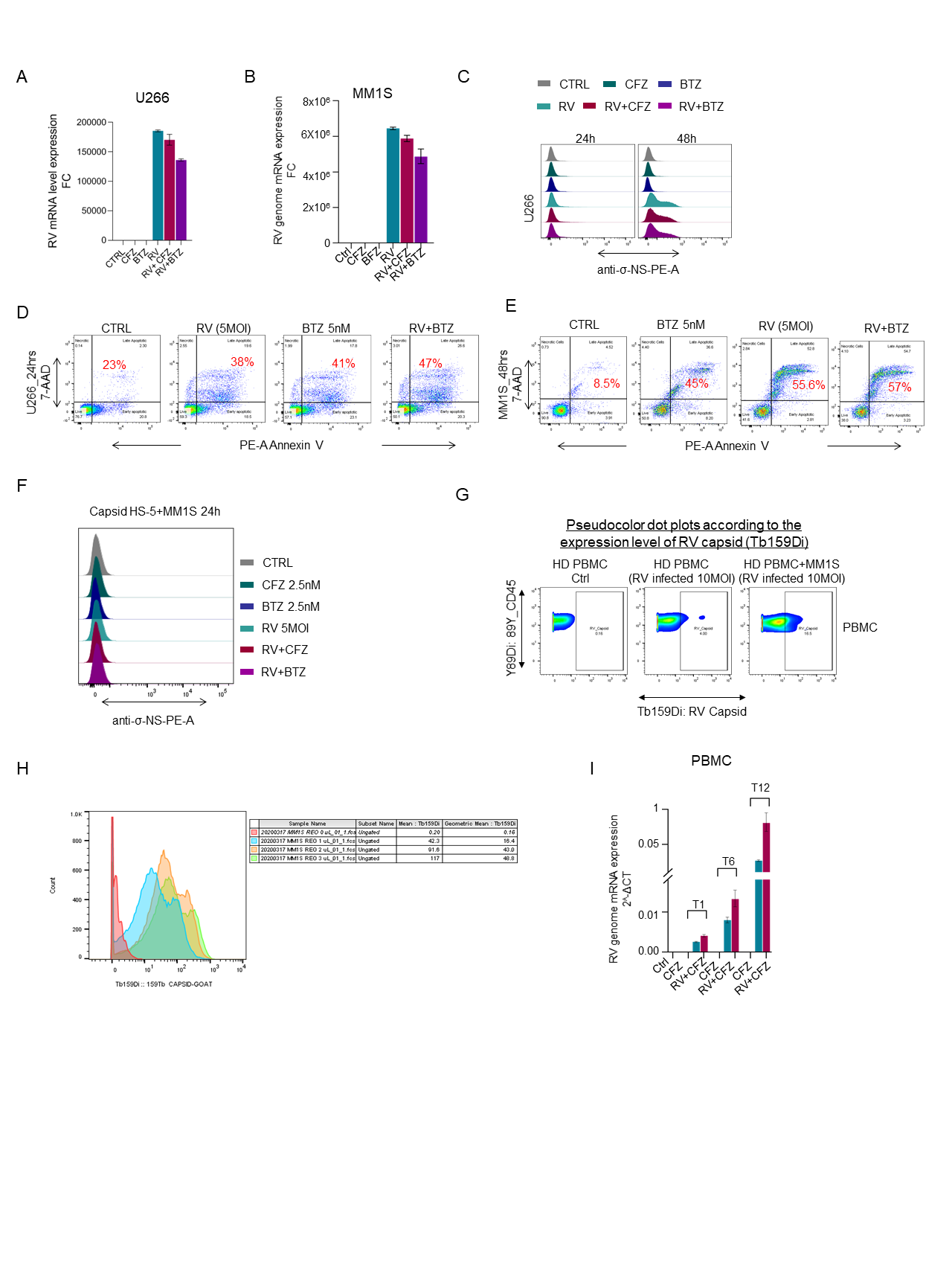

**Supplementary Figure 1. PIs potentiate RV-induced MM cell killing only with the involvement of the microenvironment**

**A-B)** MM cell lines (MM1.S and U266) were treated with CFZ or BTZ (2.5 nM) and RV (5 MOI) alone or in combination for 24hrs to assess the RV genome expression by q-RT-PCR; **C)** Offset histograms showing sigma non-structural capsid protein (σ-NS) on U266 cells treated with CFZ or BTZ (5 nM) and infected with RV (5 MOI) for 24 and 48hrs; **D-E)** Flow cytometry Annexin V dot plots showing the apoptotic rate (%) of U266 and MM1.S cells exposed to BTZ (2.5 nM) and RV (5 MOI) alone or in combination for 24 or 48hrs; **F)** BM stromal cells HS-5 were treated with CFZ or BTZ (2.5 nM) and RV (5 MOI) alone or in combination overnight, then co-cultured with MM1.S to assess RV sigma non-structural capsid protein (σ-NS) by flow cytometry; **G)** Pseudocolor dot plots according to the expression level of RV capsid (σ-NS) after CyTOF analysis of HD PBMCs infected or not with RV (10 MOI) and with or without co-culture of pre-infected MM cells as a positive control; **H)** Overlay histograms showing RV metal-conjugated titrations to assess the specificity for CyTOF staining; **I)** q-RT-PCR for the viral genome expression at different time points (0-1-6-12hrs) of PBMCs isolated from HDs.

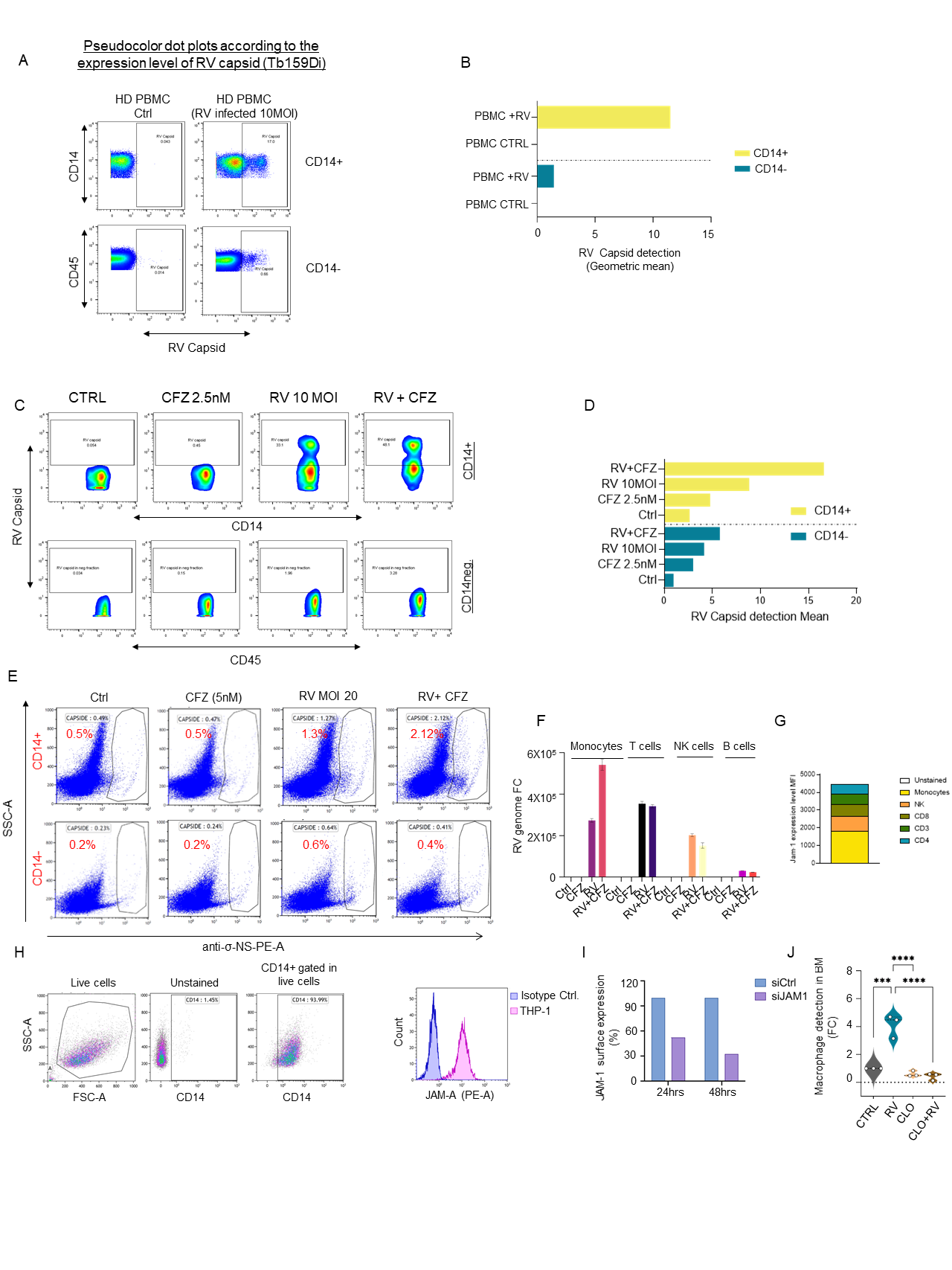

**Supplementary Figure 2. Proteosome inhibitor-enhanced viral replication requires monocytes**

**A-B)** Mass cytometry pseudocolor dot plots according to the expression level of RV capsid (Tb159Di) of HD-PBMCs infected or not with RV (10 MOI) for 24hrs (A) and bar graph showing geometric mean of RV capsid detection in CD14- and CD14+ gated sub-populations; **C-D)** Mass cytometry pseudocolor dot plots according to the expression level of RV capsid (Tb159Di) of HD-PBMCs infected or not with RV (10 MOI) alone or in combination with (CFZ 2.5nM) for 24hrs, and bar graph (D) showing geometric mean of RV capsid detection in CD14- and CD14+ gated sub-populations; **E)** Flow cytometry dot plots showing expression level of RV capsid in the same experimental conditions showing the same trend; **F)** q-RT-PCR for the viral genome expression of monocytes, T cells, NK cells and B cells isolated from the blood of an HD after 24hrs of treatment with CFZ (2.5 nM) and/or RV (5MOI); **G)** Grouped summary graph showing JAM-1 flow cytometry detection in different immune subsets (CD14+, CD56+, CD8+, CD4+, CD3+); **H)** Flow cytometry overlay and gating strategy showing JAM-1 positivity in THP-1 cell lines; **I)** Flow cytometry bar graph showing specific JAM-1 knockdown after 24 and 48hrs in THP-1 cell line; **J)** Violin plots showing bone marrow macrophage depletion of treated mice after clodronate infusion analyzed by flow cytometry (∗∗∗∗p≤ 0.0001, ∗∗∗p≤0.001).

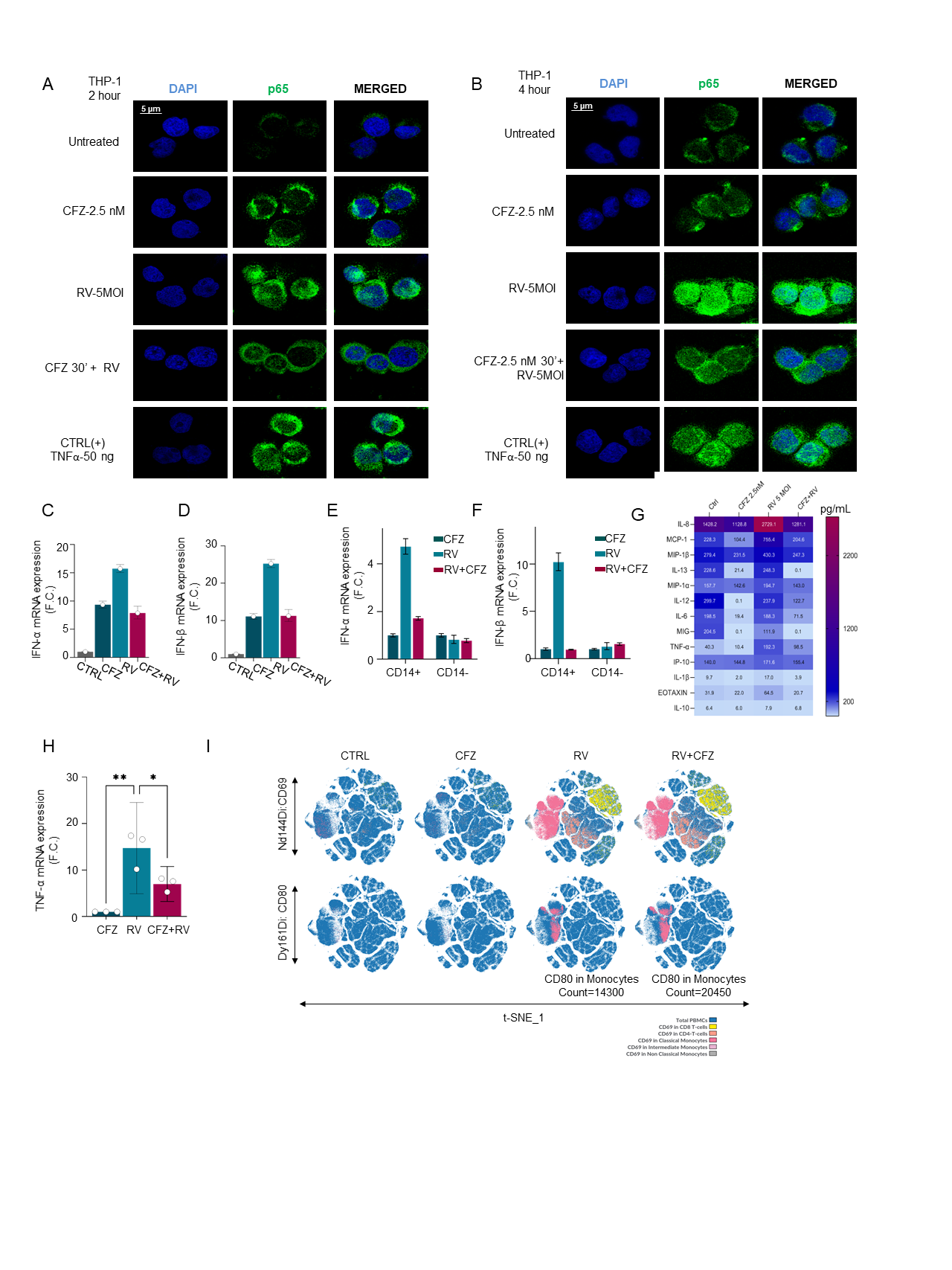

**Supplementary Figure 3. CFZ stimulates immune activation but impairs the monocyte-mediated antiviral response**

**A-B)** Representative immunofluorescence fields showing p65 staining (green) as indicated and stained with DAPI for nucleic acids (blue) in THP-1 cells treated with CFZ for 30 min then infected or not with RV (5 MOI) for 2 or 4hrs. TNF-α (50 ng/ml) was used as internal control showing p65 nuclear translocation; **C-D)** q-RT-PCR showing IFNs-type I (IFN-α and IFN-β) induction after 24hrs of RV (5 MOI) and CFZ (2.5 nM) treatments in n=3 HD PBMCs. Data are normalized to control GAPDH and expressed as mean ± s.d.in F.C. compared to the control; **E-F)** q-RT-PCR showing IFNs-type I (IFN-α and IFN-β) induction after 24hrs of RV (5 MOI) and CFZ (2.5 nM) treatments in CD14+ and CD14neg HD PBMCs. Data are normalized to control GAPDH and expressed as mean ± s.d.in F.C. compared to the control; **G)** Heatmap of multiplex cytokine profile performed on supernatant from HD-PBMC cells treated for 4hrs with CFZ, RV or both, showing 13 out 22 of the analyzed cytokines; **H)** Validation of TNF-α levels after 24hrs of treatment, n=3 biological systems data, mean ± s.d is shown; **I)** CyTOF high-fidelity FlowSOM in HD-PBMCs infected or not with RV (10 MOI) alone or in combination with CFZ (2.5 nM) showing CD69 and CD80 detection in monocyte and T cell sub-compartments.

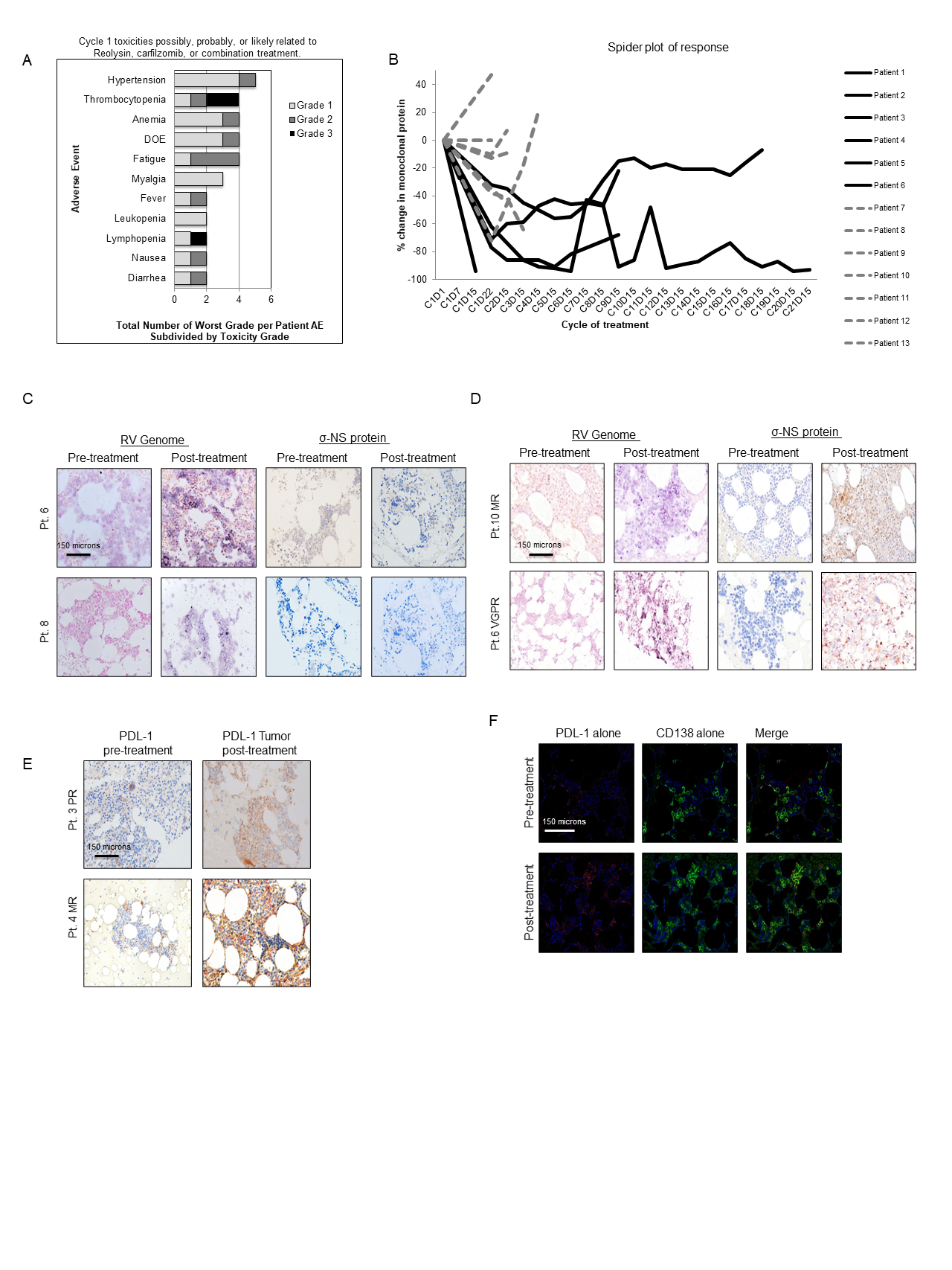

**Supplementary Figure 4. RV combined with CFZ increases viral replication in the bone marrow of MM patients**

**A)** Cycle 1 toxicities possibly, likely, or definitely due to Reolysin, carfilzomib, or the combination. The figure illustrates the total number of worst grade adverse events (AEs) per patient, subdivided by toxicity grade. The most common AE was hypertension followed by thrombocytopenia, anemia, dyspnea on exertion, fatigue, and malaise. Three patients experienced grade 3 events including two with thrombocytopenia and one with lymphopenia. No grade 4 events were reported. Abbreviation: DOE = dyspnea on exertion; **B)** Spider plot illustrating duration and depth of response in all patients treated with combinatorial Reolysin and carfilzomib. Patients treated at dose level 1 are indicated by the solid dark line and those treated at dose level -1 are indicated by the dashed gray line. Those patients treated with higher doses had deeper and more prolonged responses. Abbreviations: % = percentage, C = cycle, D = day. **C-D)** IHC in decreased magnification showing the in situ data for the detection of reoviral RNA (signal blue with pink counterstain) and reoviral capsid protein (signal brown with blue counterstain) pre- and post-treatment.; **E)** IHC detection in decreased magnification of PD-L1 protein (signal brown with blue counterstain) pre- and post-treatment. Note the strong increase in PD-L1 expression post-treatment; **F)** IF in decreased magnification showing the co-expression of PD-L1 (fluorescent red) and CD138 (fluorescent green). Note the much stronger expression of PD-L1 post treatment and that many of the cells expressing PD-L1 are myeloma cells (merged image with coexpression seen as fluorescent yellow) (scale bars at 150 micrometers).

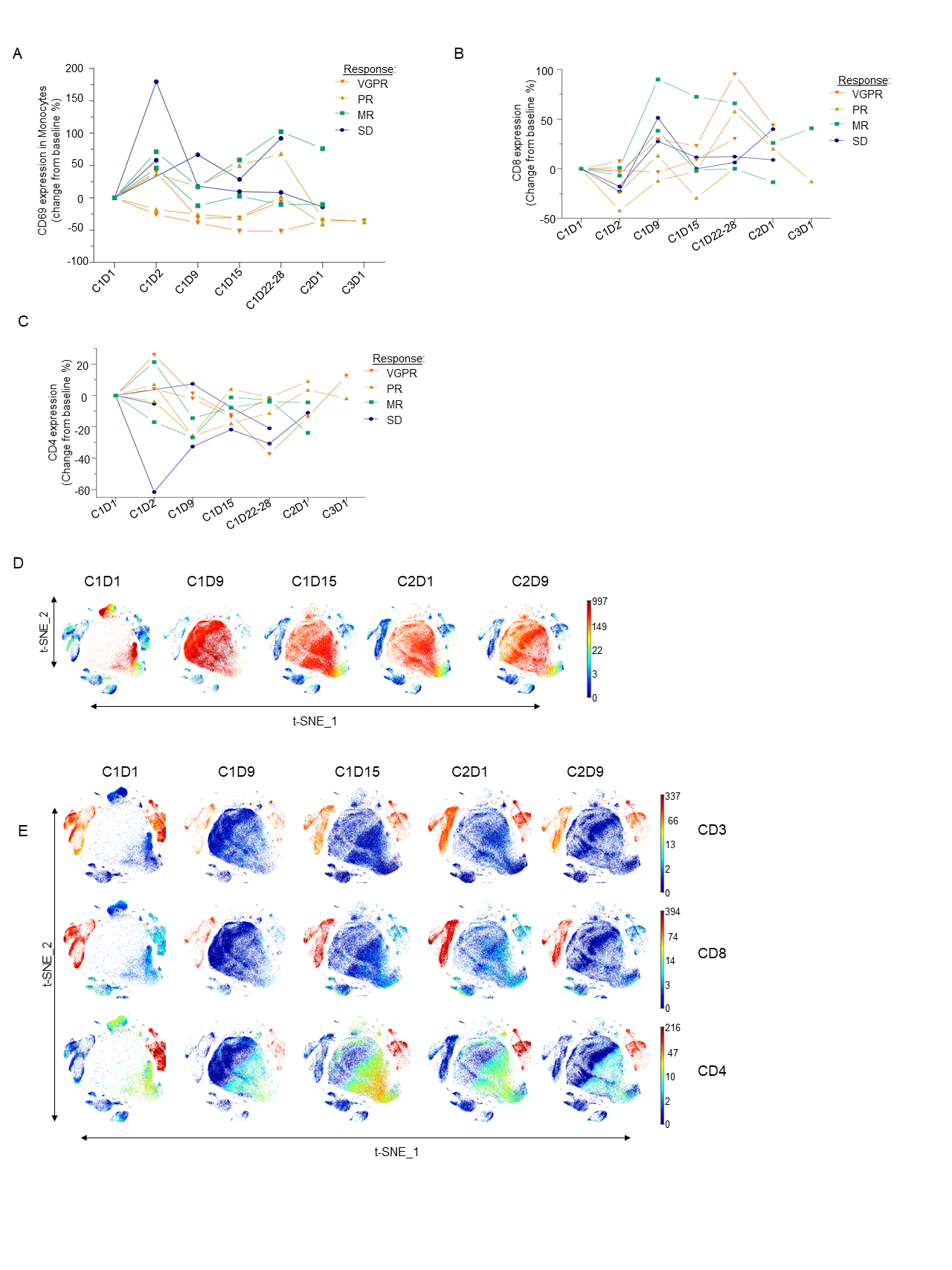

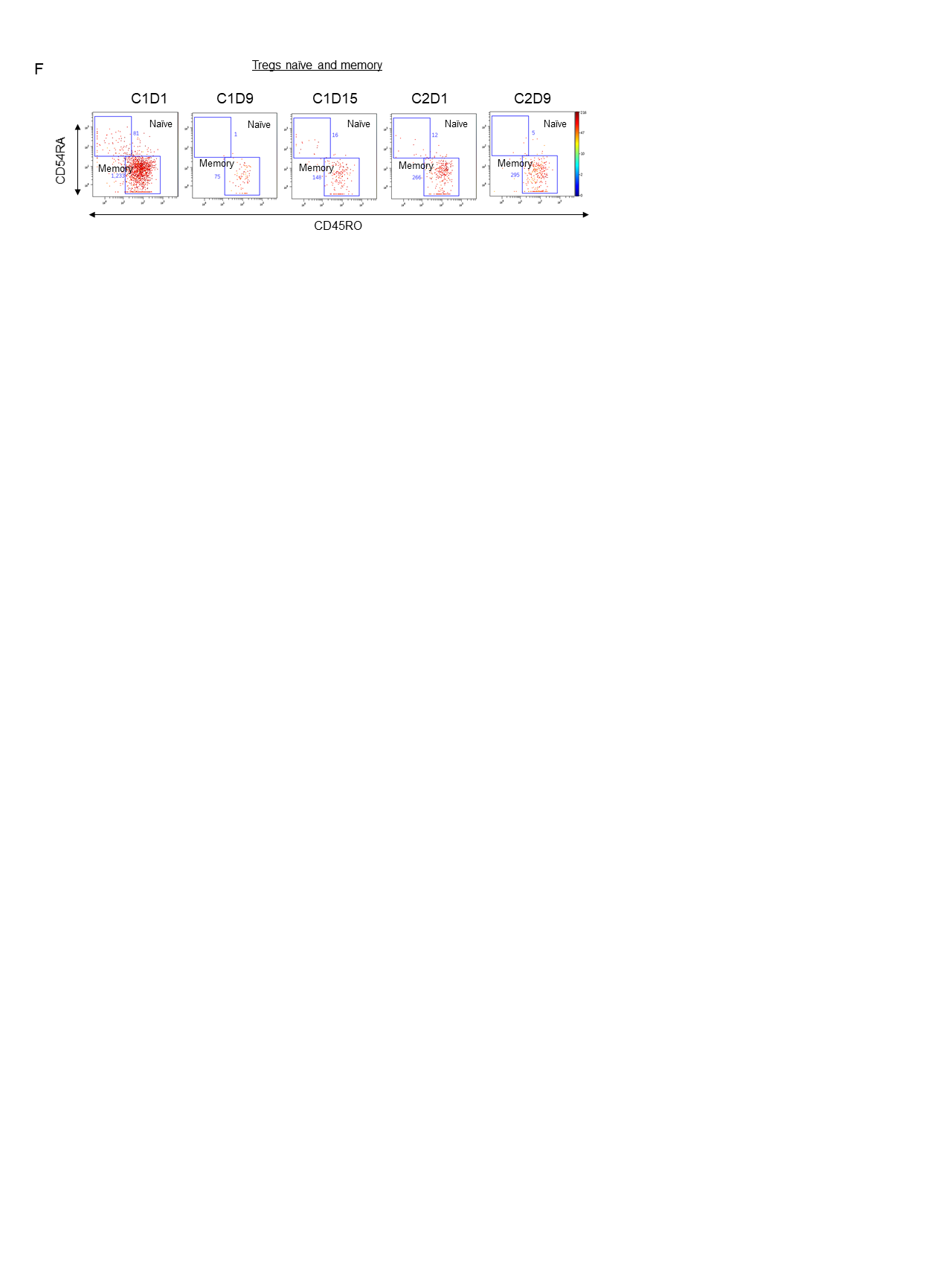

**Supplementary Figure 5. RV combined with PI activates monocytes, expands T cells, and suppresses regulatory T cells**

**A)** Line graphs representing longitudinal multiparametric flow cytometry studies on PB from relapsed MM patients enrolled in RV+CFZ Phase 1b clinical trial, showing increased CD69 activation marker in the total monocytes; Wilcoxon signed rank p-values: C1D2 > C1D1 p-value= 0.054, **B)** Line graph representing longitudinal multiparametric flow cytometry studies on PB from relapsed MM patients enrolled in the trial, showing increase in CD8 expression after treatment. Data are expressed as change from baseline %, Wilcoxon signed rank p-values: C1D9 > C1D1 p-value = 0.03; C1D22-28 > C1D1 p-value = 0.01; C2D1 > C1D1 p-value = 0.05; C1D2 < C1D1 p-value = 0.04; **C)** Line graph representing longitudinal multiparametric flow cytometry studies on the same set of patients showing CD4 expression after treatment. Data are expressed as change from baseline % and Wilcoxon signed rank p-values: C1D9 < C1D1 p-value = 0.03; C1D15 < C1D1 p-value = 0.01; C1D22-28 < C1D1 p-value = 0.007; **D)** t-SNE heatmap longitudinally displaying overall CD14 expansion of patients on trial; **E)** t-SNE heatmap showing overall expression of CD3+, CD4+, and CD8+ T cells; **F)** Dot-plots colored by channel for the signal of naïve and memory Tregs during the course of the treatment.

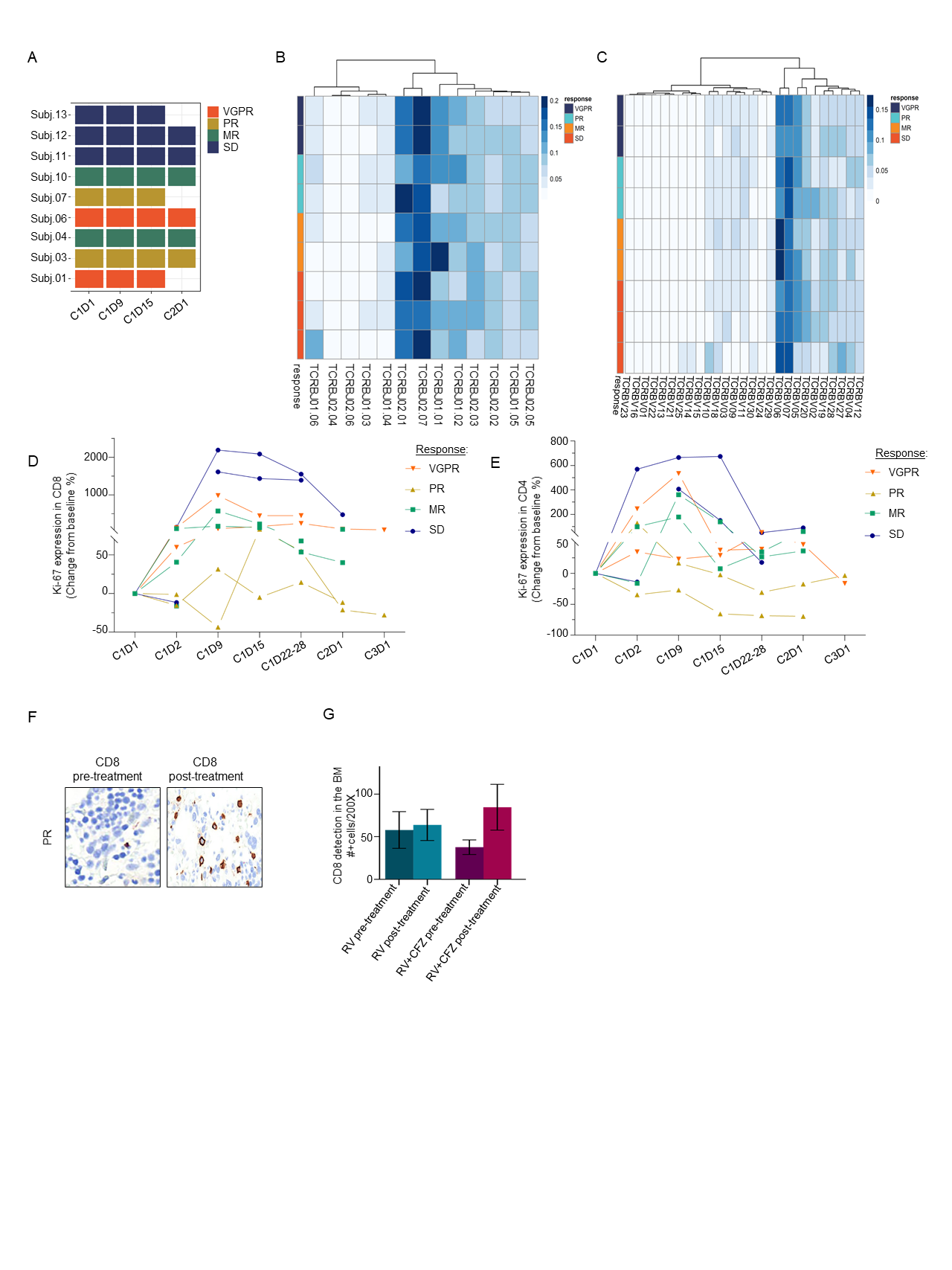

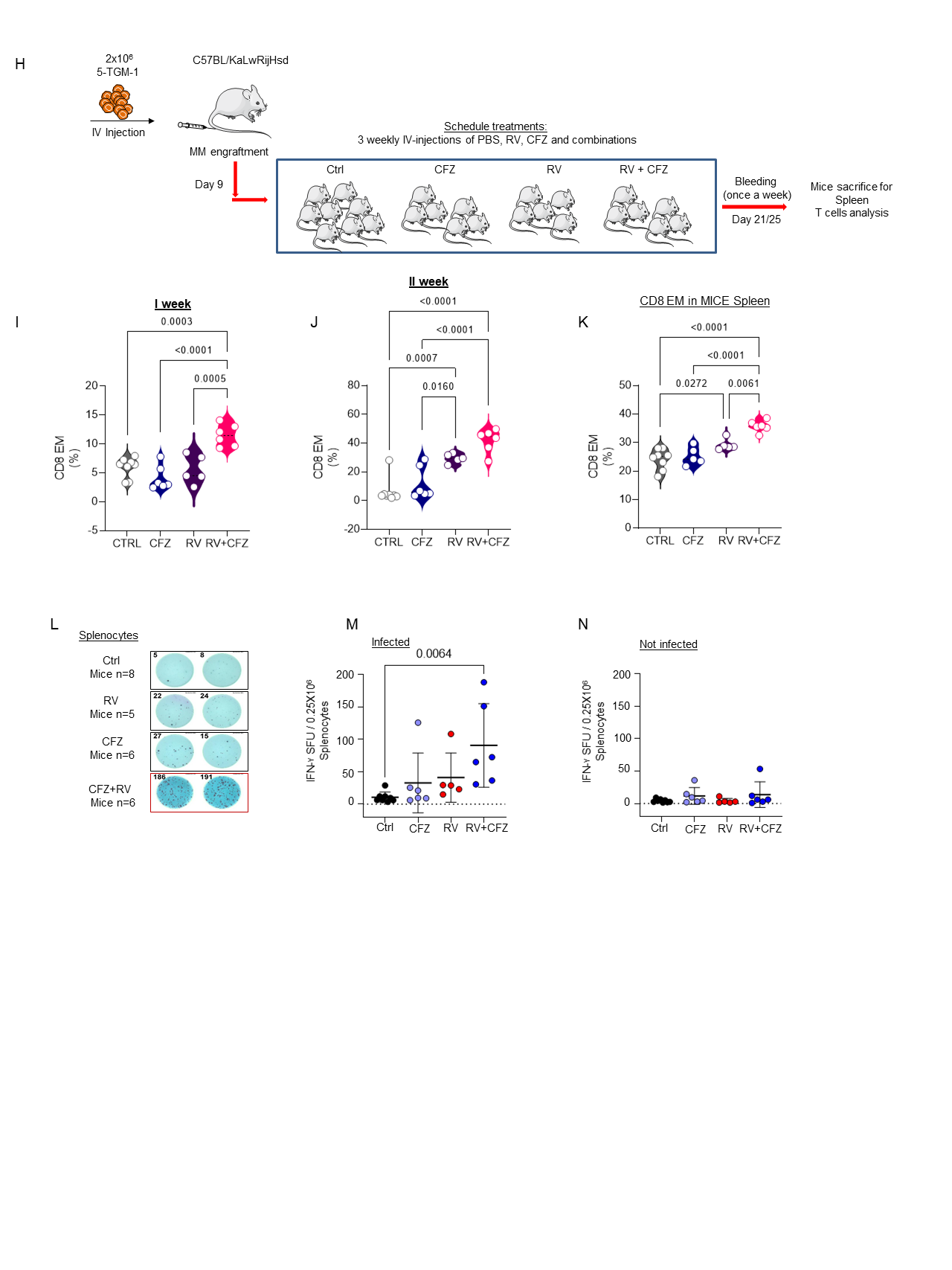

**Supplementary Figure 6. RV combined with CFZ treatment promotes T cell diversity in MM patients**

**A)** Schematic representation of an exploratory T cell repertoire study in 33 samples from 9 subjects at C1D1, C1D9, C1D15 and C2D1 with the following outcomes: VGPR (2), PR (2), MR (2) SD (3); **B-C)** Heatmaps for the V and J gene usage at the C1D1 time point showing no major differences among the subjects; **D-E)** Line graphs representing longitudinal multiparametric flow cytometry studies on PB from relapsed MM patients enrolled in RV+CFZ Phase 1b clinical trial showing Ki-67 expression in CD8 (D), Wilcoxon signed rank p-values: C1D9 > C1D1 p-value = 0.02; C1D15 >C1D1 p-value = 0.007; C1D22-28 > C1D1 p-value = 0.004; Ki-67 expression in CD4 (E) and Wilcoxon signed rank p-values: C1D9 > C1D1 p-value = 0.01; C1D22-28 ≠ C1D1 p-value = 0.54; **F-G)** IHC detection of CD8 protein (signal brown with blue counterstain) pre and post treatment in a patient with PR (F) and bar graph in 5 patients following RV monotherapy and combinatorial RV+CFZ. Each value represents the number of positive cells per 200x field. Significant increases were noted in CD8+ T cells following treatment with dose level – 1 combination treatment. p=0.060 (G); **H)** Schematic representation of mice experiment. C57BL/KaLwRij mice were injected IV with 2x10^6^ 5TGM1 cells, then randomized in four treatment groups. Mice were treated once a week with intravenous injection of RV alone (2×10^7^ PFU) (n=5) or biweekly with intraperitoneal injection of CFZ (1.6 mg/Kg) (n=6), or the combination of both (n=6). Diluent (PBS 1X) treated mice were also used as control (n=8); **I-J)** Blood of mice was collected every week to assess circulating CD8+ EM T cells. Violin plot showing CD8+ EM (%) after the first (I) and the second (J) week of treatment; **K)** Violin plot showing CD8+ EM T cells in harvested spleen after the treatment; **L-M-N)** Representative IFN-ƴ ELISPOT assay showing responses of splenocytes isolated from treated mice and cocultured *ex vivo* with 5TGM-1 cells infected or not with RV (5MOI) (L); bar graphs showing numbers of IFN-ƴ spot forming units (SFUs) after stimulation of splenocytes with 5TGM-1 cells infected (M) or not (N) with RV (5MOI).

**Supplemental methods**

*Primary samples*

Primary samples (total BM aspirates and peripheral blood samples) from MM patients and healthy donors (HDs) were obtained from The Ohio State University and City of Hope Leukemia Tissue Banks. Healthy donor samples were obtained from the City of Hope hematopoietic tissue repository (IRB#16352). Specifically, the cellular fraction of the PBMCs was isolated using Ficoll-Paque Plus (GE, Healthcare, Life Science) following the manufacturer’s instructions.

*Cell culture*

MM cell lines (MM.1S, NCI-H929, U266, and RPMI-8226), THP-1 MΦ cell line and the BM stromal cell line HS-5 were purchased from ATCC. The GFP+/Luc+ MM.1S stable line was a gift from Irene Ghobrial (Dana-Farber Cancer Institute, Boston, Massachusetts, USA). L363, an MM cell line, was provided by Jonathan J. Keats (https://www.keatslab.org/). All the cell lines were cultured in RPMI-1640 medium supplemented with 10% fetal bovine serum (FBS) (Cat. #019K8420, Sigma), 100 IU/mL penicillin, and 100 μg/mL streptomycin (Cat.#15140-122, Gibco).

*Immunoblotting*

CD14+ purified cells from healthy donors were lysed in RIPA buffer (89901, Thermo Scientific), supplemented with protease and phosphatase inhibitors, then sonicated (30” Pulse ON 02” Pulse OFF 03” 60% amplification). Lysates were then clarified by spinning at 14,000 rpm at 4^°^C and protein concentration quantified by BCA Protein Assay (23227, Thermo Scientific). Forty micrograms of proteins were denatured in boiling SDS sample buffer, resolved on 4%-20% gradient gels (Cat.# 5671093, Bio-Rad), and transferred to nitrocellulose membranes (Cat.# 1704271 Bio-Rad). After blocking nonspecific binding of antibody with 5% BSA (Fisher BioReagents), blots were probed with one of the following antibodies: anti-RV σ-NS (1:1000; provided by Oncolytics Biotech Inc.) to assess viral replication for σ1 protein; GAPDH (Cat.# sc-32233, Santa Cruz Biotechnology) was used as internal control. Blots were washed three times for 15 minutes with TBST 1X, and stained with horseradish peroxidase (HRP)-conjugated secondary antibodies (diluted 1:4000) for 2hrs at room temperature. Primary antibodies were detected by binding donkey anti-goat IgG (H+L) (Cat.# A16005, Invitrogen) and goat anti-mouse IgG-HRP (NA931, GE Healthcare), and using an enhanced chemiluminescent visualization system (Cat.#RPN2209 ECL Western Blotting Detection Reagents, GE Healthcare). Primary and secondary antibodies were diluted according to the manufacturer instructions. The bands were quantified by densitometry analyses using Image Lab program (Biorad) and normalized to GAPDH.

*Flow cytometry -- surface staining*

For surface expression analysis in MM cell HD PBMCs or isolated monocytes and primary samples, cells were washed with PBS 1X and stained for 30 minutes in staining solution (PBS+2%FBS) using antibodies listed in **Supplemental Table 6**, to determine median of fluorescence and percentage of expression in the different cell populations. Cells were washed and immediately analyzed on LSRII (Becton Dickinson) or Fortessa X-20 (Becton Dickinson). Analysis was conducted using FlowJo™ Software (version 10.7.1).

*Flow Cytometry -- intracellular detection*

MM lines cells, HD PBMCs or isolated monocytes, and primary samples were washed with PBS and fixed with 1.6% formaldehyde solution (Ref. #28908, Thermo Scientific) for 15 minutes at room temperature. Cells were then washed with perm/wash buffer (Cat. # 51-2091KZ) and stained for cytoplasmic RV detection with anti-RV σ-NS (1:1000; provided by Oncolytics Biotech Inc.) diluted in 100 µL of perm/wash buffer for 30 minutes at room temperature. After incubation, cells were washed with 2 mL of perm/wash buffer and stained with secondary antibody Goat IgG (H+L) PE-conjugated antibody (Cat. # F0107 R&D Systems) and diluted in 100 µL of perm/wash buffer for 30 minutes at room temperature. After secondary antibody incubation, cells were finally washed with PBS and immediately analyzed on LSRII (Becton Dickinson) or Fortessa X-20 (Becton Dickinson). For Ki 67 detection, Ki-67 AF700 Cat. #561277, was used, and for Granzyme B the PE conjugated Cat. #561142 was used, both from BD Biosciences. Analysis was conducted using FlowJo™ Software (version 10.7.1). Untreated and unstained cells were used as controls.

*Flow cytometry based-killing assay ex-vivo*

Effector cells, PBMCs isolated from HDs, and target MM.1S GFP+/Luc+ cells were co-cultured at different ratio 8:1 for 12hrs. Briefly, PBMCs (2x10^6^ cells/well for the 8:1 ratio) were seeded at the appropriate concentration, then retreated for 30 min in the presence or absence of CFZ, then infected overnight with RV (5 MOI). After incubation, cells were washed and co-cultured with the appropriate ratio of target cells (2.5x10^5^ cells/well for the 8:1 ratio) for 12hrs. After incubation, the killing induction (%) was assessed by flow cytometry using 7-aminoactinomycin D (7-AAD) (Cat. #51-68981E BD Biosciences). Data are expressed as the mean ± SEM (n=3). The experiment was conducted in two independent biological systems.

*RNA isolation and analysis*

Total cellular RNA was extracted by using TRIZOL reagent (Cat. #15596018 Invitrogen Corporation) and RNA Clean-Up and Concentration Kit (Cat. #43200 Norgen) according to the manufacturer’s protocols. cDNA synthesis was performed by using the High Capacity cDNA Reverse Transcription Kit (Applied Biosystems, Cat# 4368814). Reverse transcription reactions were run using a Mastercycler pro. Quantitative real time-PCR (qRT-PCR) was performed with the TaqMan method (Applied Biosystems), according to the manufacturer’s instructions. The appropriate TaqMan probes for mRNA quantification were purchased from Applied Biosystems, and all reactions were performed in triplicate. The following probes were used: (HS99999905_m1) GAPDH used as endogenous control; (HS00989291_m1) INF-γ; (Hs00961622 m1) IL-10; (Hs00174131 m1) IL-6; (Hs01077958 S1) IFNB1; (Hs00174128 m1) TNFα; (Hs00265051 S1) IFNA2.

For the quantification of viral RNA genome extracted from infected HD PBMCs and MM cell lines q-RT-PCR reactions were conducted using the PowerUp SYBR Green Master Mix (Applied Biosystems Cat.# 4367659) according to the manufacturer’s instructions and the following primers: Reo9: 5′-TG CGC AAG AGG CAG CAA TCG-3′ and Reo10: 5′-TT CGC GGG CCT CGC ACA TTC-3′; GAPDH FD: 5′- CTG CAC CAC CAA CTG CTT -3′ and GAPDH RV: 5′- CAT GAC GGC AGG TCA GGT -3′.

*Gene silencing*

THP-1 cells were transfected by electroporation using the Nucleofector4D system (Lonza). Briefly, on day 1, two batches of 6×10^6^ cells were each resuspended in 100 μL of the nucleofector SF solution containing 50 pmol siJAM1 (Herizon Discovery, catalog M-005053-01-0010) or 1 µg pmax GFP Vector (Lonza; V4XC-2024) and transferred to a cuvette. After electroporation, cells were kept in culture in 10 mL of complete growth media. Program FF-100 was used for THP1 cells. On day 3 (48hrs post-transfection), THP1 (siCtrl or siJAM1) cells were transfected with Reovirus (5 MOI) alone or in combination with CFZ (2.5 nM) for 24hrs. On day 4, Cells were harvested for further experiment.

*Immunofluorescence and confocal microscopy*

Jurkat and THP-1 cells (1 million/well) were seeded into 12 well plates and were treated with RV, CFZ and TNF-α individually or in combinations. Post-treatment, cells were collected, washed with 1X PBS and cyto-spinned on \to glass slides. The cells were then fixed in 4% paraformaldehyde for 30 min at room temperature (RT), washed (3 times in 1X PBS), and then permeabilized by 0.1% Triton-X 100 for five minutes at RT, followed by washing (3 times in 1X PBS). The cells were then blocked in blocking buffer (3% FBS in 1X PBS) for 1hr at RT and washed (3 times in 1X PBS). The cells were subsequently incubated with primary antibody Rabbit NF-KappaB-p65 (CST – Cat. #8242) 1:700 overnight at 4°C. Post-incubation, cells were washed (3 times in 1X PBS) and incubated in secondary antibody Alexa Fluor 488 Donkey Anti-Rabbit IgG Invitrogen Cat# - A21206 1:500. After incubation with the secondary antibody at RT for 1hr, cells were washed (3 times in 1X PBS) and mounted in Fluoroshield with DAPI (Cat: Qs4-20ML) from enQuire BioReagents. Slides were observed under Confocal LSM 880 microscope.

*Statistical Analyses of TCR-β sequencing results*

Simpson Clonality was calculated on productive rearrangements by: $\sqrt{\sum p_{i}^{2}}$ where *p_i_* is the proportional abundance of rearrangement *I*, and *N* is the total number of rearrangements. Clonality values range from 0 to 1 and describe the shape of the frequency distribution: clonality values approaching 0 indicate a very even distribution of frequencies, whereas values approaching 1 indicate an increasingly asymmetric distribution in which a few clones are present at high frequencies.

*Immunohistochemistry*

An antibody against reovirus capsid protein was provided by Dr. Matt Coffey (Oncolytics Biotech Inc.). The antibodies against PD-L1, CD138, Caspase 3, and CD8 were obtained from Abcam (Cambridge, MA). The viral RNA in situ hybridization protocol has been previously described [16] and used digoxigenin-tagged locked nucleic acid probes from Exiqon. The cell counts for CD8, PD-L1, Caspase 3, and reovirus RNA/protein were compiled by counting the number of positive cells in multiple 200x fields. At least 3000 cells were counted, and mean (and standard deviation) was derived and analyzed with the InStat Statistical Analysis Software (version 3.36).

*Cytokines array*

Cytokines array was performed using a Custom Luminex Bead Reassignment Product (CUSTOM-LXSA-H-25) purchased from Bio-Techne. We analyzed the following set of 22 cytokines: CCL2/JE/MCP-1(BR25); CCL3/MIP-1alpha (BR35); CCL4/MIP-1 BETA (BR47); CCL11/Eotaxin (BR77); CD25/IL-2R alpha (BR47); CXCL9/MIG (BR52); GM-CSF (BR46); IFN-alpha (BR63); IFN-beta (BR21); IFN-gamma (BR29); IL-1 alpha/IL-1F1 (BR38); IL-1 beta/IL-1F2 (BR28); IL-2 (BR43); IL-4 (BR39); IL-5 (BR53); IL-6 (BR13); IL-7 (BR20); IL-8/CXCL8 (BR18); IL-10 (BR22); IL-12/IL-23 p40 (BR67); IL-13 (BR64); IL-17/IL-17A (BR42); TNF-alpha (BR12).

*ELISpot assay*

The assay was performed on splenocytes isolated from mice infected, or not, ex-vivo with RV (5MOI) treated with RV and CFZ alone or in combination. Specifically, 2x10^6^ 5-TGM1 cells were intravenously injected in syngeneic C57BL/KaLwRij mice, after 9 days from the injection, mice were randomly divided in four treatment groups. Mice were treated once a week with intravenous injection of RV alone (2×10^7^ PFU) (n=5), biweekly with intraperitoneal injection of CFZ (1.6mg/kg) (n=6), or the combination of both (n=6). Diluent (PBS 1X) treatment of mice was used as control (n=8). For ELISpot assay all reagents used were filtered through a 0.22-micron filter. Wells of 96-well Multiscreen HTS Plates (Millipore, Billerica, MA) were pre-wet with 35% methanol and coated with 100 μl primary anti-IFN-γ antibody solution (10 μg/ml of clone AN18 from Mabtech in PBS, pH 7.4) and incubated overnight at 4°C. After washing with PBST (PBS-Tween 0.02% solution) and PBS, nonspecific antigen binding was blocked with 200 μl of RPMI media containing 10% (v/v) FBS for 2 hours at room temperature. Following the blockade, 0.25 × 10^6^ cells from mouse splenocytes in 100 μl of RPMI were co-cultured with 0.25 × 10^6^ 5-TGM1 cells (infected with 10 MOI reovirus or uninfected) and added to each well. After incubation for 48 hours in humidified 5% CO_2_ at 37°C, cells were removed by washing, and 100 μl of biotinylated secondary anti-IFN-γ antibody (clone R4-6A2, Mabtech in blocking buffer) was added to each well. Following a 2-hour incubation and washing, HRP-conjugated streptavidin was diluted 1:1000, and wells were incubated with 100 μl for 1 hour at room temperature. Following washing, wells were incubated for 30 min at room temperature with 100 μl of TMB detection reagent, and spots were counted with an automated ELISpot Reader System (Cellular Technology Limited, C.T.L).
